# Transcriptomic analysis of frontotemporal lobar degeneration with TDP-43 pathology reveals cellular alterations across multiple brain regions

**DOI:** 10.1101/2021.10.06.21264635

**Authors:** Rahat Hasan, Jack Humphrey, Conceição Bettencourt, NYGC ALS Consortium, Tammaryn Lashley, Pietro Fratta, Towfique Raj

## Abstract

Frontotemporal lobar degeneration (FTLD) is a group of heterogeneous neurodegenerative disorders affecting the frontal and temporal lobes of the brain. Nuclear loss and cytoplasmic aggregation of the RNA-binding protein TDP-43 represents the major FTLD pathology, known as FTLD-TDP. To date, there is no effective treatment for FTLD-TDP due to an incomplete understanding of the molecular mechanisms underlying disease development. Here we compared post-mortem tissue RNA-seq transcriptomes from the frontal cortex, temporal cortex and cerebellum between 28 controls and 30 FTLD-TDP patients to profile changes in cell-type composition, gene expression and transcript usage. We observed downregulation of neuronal markers in all three regions of the brain, accompanied by upregulation of microglia, astrocytes, and oligodendrocytes, as well as endothelial cells and pericytes, suggesting shifts in both immune activation and within the vasculature. We validate our estimates of neuronal loss using neuropathological atrophy scores and show that neuronal loss in the cortex can be mainly attributed to excitatory neurons, and that increases in microglial and endothelial cell expression are highly correlated with neuronal loss. All our analyses identified a strong involvement of the cerebellum in the neurodegenerative process of FTLD-TDP. Altogether, our data provides a detailed landscape of gene expression alterations to help unravel relevant disease mechanisms in FTLD.

## Introduction

Frontotemporal dementia (FTD) is a spectrum of neurodegenerative disorders affecting the frontal and anterior temporal lobes of the brain, manifesting clinically as disturbances in behaviour or language. The neuropathological correlate of FTD, Frontotemporal Lobar Degeneration (FTLD), can be subclassified into three distinct subgroups based on specific protein inclusions. Around 50% of patients present with cytoplasmic inclusions of transactive response DNA-binding protein 43 kDa (TDP-43), leading to a histopathological diagnosis of FTLD-TDP (Neumann et al. 2006). TDP-43 pathology is present in a range of other neurodegenerative diseases, most notably amyotrophic lateral sclerosis (ALS) (Neumann et al. 2006). Approximately 40% of FTD patients have a positive family history of disease (Goldman et al. 2005). Mutations in two genes, *C9orf72* and *GRN*, explain the majority of familial FTLD-TDP (Renton et al. 2011; Baker et al. 2006; DeJesus-Hernandez et al. 2011; Cruts et al. 2006), but several other causal genes have been identified that explain a smaller fraction of cases. These genes include *TARDBP*, the gene encoding TDP-43, as well as *VCP, SQSTM1*, and *TBK1* (Borroni et al. 2009; Bersano et al. 2009; Le Ber et al. 2013; Gijselinck et al. 2015).

Much research has focused on understanding the formation and consequences of TDP-43 aggregation in FTLD. TDP-43 is an RNA binding protein that regulates many aspects of mRNA processing, including alternative splicing and polyadenylation (Polymenidou et al. 2011; Tollervey et al. 2011; Rot et al. 2017). Under conditions not currently understood, TDP-43 is mislocalized from the nucleus to the cytoplasm, where it undergoes post-translational modifications such as hyperphosphorylation, ubiquitination, and N-terminal truncation (Neumann et al. 2006). Both nuclear loss of function (LOF) and cytoplasmic gain of function (GOF) mechanisms have been proposed for the consequences of TDP-43 mislocalization (Ling, Polymenidou, and Cleveland 2013). Supporting a potential role for LOF in FTLD, recent studies have demonstrated that loss of nuclear TDP-43 alters the expression and splicing of its target mRNAs, many of which are involved in synapse organization and plasticity (Polymenidou et al. 2011; Honda et al. 2013). Nuclear loss of TDP-43 leads to aberrant cryptic splicing of these targets, including key neuronal genes *STMN2* and *UNC13A* (Klim et al. 2019; Melamed et al. 2019; Brown et al. 2021; Ma et al. 2021). Thus, TDP-43 nuclear loss may play a role in the neurodegenerative process of FTLD.

FTLD is characterised by neuronal loss and an inflammatory response driven by glial cells. Both excitatory glutamatergic pyramidal cells and inhibitory GABAergic neurons are lost or dysregulated in post-mortem FTLD-TDP brains (Hughes et al. 2018; Šarac et al. 2008; Murley et al. 2020; Ferrer 1999). A unique subgroup of excitatory projection neurons known as von Economo neurons (VENs) has been shown to be selectively vulnerable to TDP-43 pathology, dying particularly early in disease progression (Santillo, Nilsson, and Englund 2013; Nana et al. 2019; Gami-Patel et al. 2019). Although the cerebellum is spared from TDP-43 pathology, it is an open question whether neurodegeneration also occurs there in FTLD. In cases caused by *C9orf72* mutations, repeat-associated non-ATG (RAN) translation leads to the accumulation of *C9orf72* dipeptide repeat protein, which may have neurotoxic effects on the cerebellum (Yousef et al. 2017; Gendron et al. 2015). Microglia and astrocytes are essential for clearing of debris and maintaining brain homeostasis (Jung and Chung 2018). However, in response to neurodegeneration, these cells are believed to switch to an activated state, producing pro-inflammatory cytokines, chemokines, and reactive oxygen species that contribute to a state of neuroinflammation (Glass et al. 2010; Ramesh, MacLean, and Philipp 2013; Liddelow et al. 2017). Although this may initially be beneficial for removing surrounding protein aggregates and dying neurons, sustained inflammatory responses can damage neurons and synapses, ultimately exacerbating neurodegeneration.

Although recent advancements have improved our understanding of the molecular basis of FTLD-TDP, it is still unclear how mutations in the different causal genes lead to disease, and what the underlying pathways are. Transcriptome profiling of human post-mortem brain tissue is necessary to gain a deeper insight into the functional pathological changes in all FTLD-TDP subtypes. To address this need, we have assembled a cohort of RNA-seq generated from post-mortem brain samples from FTLD-TDP and non-neurological disease control patients. We assessed the shared and distinct transcriptomic changes associated with FTLD-TDP across the frontal cortex, temporal cortex, and cerebellum. We performed differential gene expression, differential transcript usage, and cell-type deconvolution analysis, identifying changes in cellular composition and cellular pathways across all three brain regions.

## Methods

### NYGC ALS Consortium – FTD cohort

RNA-seq samples were obtained from the January 2020 data freeze of the New York Genome Center (NYGC) ALS Consortium. The FTD cohort is a collection of postmortem brain tissue from 41 non-neurological controls and 49 FTD donors, as provided by the UCL Queen Square Brain Bank. Among the FTD donors, 37 have TDP-43 pathology (FTLD-TDP), of which 9 have mutations in *C9orf72*, 7 in *GRN*, 2 in *TBK1*, and 19 are sporadic cases. Together, the 37 FTLD-TDP donors and 41 controls provide a total of 169 RNA-seq samples and represent the FTLD-TDP subcohort. De-identified clinical information for the cohort is presented in **Supplementary Table 1**. The NYGC ALS Consortium samples presented in this work were acquired through various institutional review board (IRB) protocols from member sites and the Target ALS postmortem tissue core and transferred to the NYGC in accordance with all applicable foreign, domestic, federal, state, and local laws and regulations for processing, sequencing, and analysis. The Biomedical Research Alliance of New York (BRANY) IRB serves as the central ethics oversight body for NYGC ALS Consortium. Ethical approval was given and is effective through 08/22/2022. Brain tissue dissection was performed by pathologists at the UCL Queen Square Brain Bank. Cortical and cerebellar regions were removed from each subject and divided into left and right hemispheres. One hemisphere was flash-frozen for transcriptome sequencing while the other was sectioned for histopathological evaluation.

The RNA sequencing procedures of the NYGC have been previously described (Tam et al. 2019). RNA was isolated from the frozen brain tissue with TRIzol reagent and purified using RNeasy mini columns (Qiagen). RNA integrity numbers (RIN) for the brain samples were estimated on a Bioanalyzer (Agilent Technologies). RNA-Seq libraries were generated starting from 500 ng of total RNA using the KAPA Stranded RNA-Seq Kit with RiboErase (KAPA Biosystems) to remove rRNA and Illumina-compatible indexes (NEXTflex RNA-Seq Barcodes, NOVA-512915, PerkinElmer, and IDT for Illumina TruSeq UD Indexes, 20022370). Pooled libraries (average insert size: 375 bp) were then sequenced on either an Illumina HiSeq 2500 (125 bp paired end) or Illumina Novaseq (100 bp paired end). The sequenced samples were then subjected to extensive quality control protocols to confirm variables such as sex, tissue, and *C9orf72* repeat expansion status. *C9orf72* repeat expansions were identified on genotyped samples using the Asuragen AmplideX PCR/CE C9orf72 Kit and ExpansionHunter. Only samples with RNA integrity number (RIN) greater than 5 were chosen for study, due to the impact of low RIN on gene expression (Schroeder et al. 2006). Of the 169 samples from the FTLD-TDP subcohort, 23 FTLD-TDP and 12 control samples did not meet the RIN cutoff.

### RNA-seq data processing and sample selection

RNA-seq samples were uniformly processed using RAPiD-nf, a processing pipeline implemented in the NextFlow framework. After trimming adapter sequences with Trimmomatic (version 0.36), the samples were aligned to the hg38 build of the human reference genome (GRCh38.primary_assembly) using STAR (2.7.a). Gene counts were generated using RSEM (1.3.1). Quality control was performed using SAMtools and Picard, modeling the criteria of the Genotype Tissue Expression Consortium (GTEx Consortium 2020). To identify outliers, we performed principal component analysis (PCA) on the voom-normalised RNA-seq expression matrix, checking for points that did not cluster with their tissue type. One outlying sample was identified and removed from the study. Repeating PCA with this sample removed did not show any additional outliers or changes in clustering. For the present study, we employed samples from the frontal cortex, temporal cortex and cerebellum, spanning 74 samples from 30 FTLD-TDP donors and 55 samples from 28 non-neurological controls. The 74 FTLD-TDP samples come from 16 sporadic cases, and 14 genetic cases of which 9 have mutations in *C9orf72*, 4 in *GRN*, and 1 in *TBK1*.

### Covariate adjustment

Before performing DGE analysis, the RNA-seq expression matrix was normalised and adjusted for covariates. Normalisation was performed using trimmed mean of M values and transformed with the limma::voom() function (Law et al. 2014). Lowly expressed genes were removed using a threshold of >1 counts per million in at least 90% of the samples. Covariate adjustment was performed separately for each brain region, and the steps described here were modeled from a large differential expression study (Fromer et al. 2016). First, clinical variables were combined with sequencing variables and technical metrics from Picard. Then, potential confounders were determined by ranking the variables based on their contributions to gene expression variance. The variance contributions of the covariates to each gene were scored using the limma::selectModel() function, which returned the number of genes with a lower Bayesian Information Criterion (BIC) as a result of adding the covariate to a base model containing only disease as the predictor (**Supplementary Fig. 1)**. For a given gene, a lower BIC indicated an improvement over the base model, or a larger portion of the variance explained by the covariate. The top ten covariates that improved the BIC for the largest number of genes were considered potential confounders. Correlating technical factors with each other allowed us to select a set of distinct factors for modelling (**Supplementary Fig. 2**).

To find the subset of covariates resulting in the best fitting linear regression model for DGE analysis, stepwise regression was performed by successively adding each variable to the base model. After evaluating each successive model with the limma::selectModel() function **(Supplementary Fig. 3a)**, we chose the model that improved the BIC for the largest number of genes. Using an orthogonal approach, variancePartition (Hoffman and Schadt 2016) was run on the covariates to quantify their contributions to gene expression variance **(Supplementary Fig. 3b)**. The following models were fitted for each brain region:

#### Selected models for FTLD-TDP vs control differentiation expression

| **Region** | **Model** |
| --- | --- |
| Frontal Cortex | *expression ~ disease + sex + age + median 3' bias* |
| Temporal Cortex | *expression ~ disease + sex + age + median 3' bias + % intronic bases + % ribosomal bases* |
| Cerebellum | *expression ~ disease + sex + age + % R2 transcript strand reads* |

### Differential gene expression analyses

Adjusting for the covariates described above, we performed differential gene expression (DGE) analysis to compare the FTLD-TDP cases with controls. The limma package was used to compute the log_2_-fold changes, t-statistics, and P-values for all genes tested in each brain region **(Supplementary Table 2)**. Genes having an adjusted P < 0.05 were considered differentially expressed. We then correlated the log_2_-fold change effect sizes of each gene tested between each pair of brain regions. Next, we repeated DGE analysis, but split the FTLD-TDP cases by *C9orf72* repeat expansion status. After comparing the *C9orf72* and non*-C9orf72* cases with controls, we correlated the log_2_-fold changes of all genes tested between each disease group in each region. DEGs (adjusted P < 0.05) were overlapped between the two groups. To check for confounding effects of post-mortem interval (PMI) on gene expression, we performed DGE analysis on the cases and controls respectively by using PMI as a continuous variable. We then associated the log_2_-fold changes of the PMI-related genes and the FTD genes in each brain region and calculated Pearson correlation coefficients to quantify the effect of PMI.

### Gene set enrichment analyses

Gene set enrichment analyses (GSEA) were conducted using the clusterProfiler package (Yu et al. 2012). The inputs included the differentially expressed genes (DEGs), ranked by t-statistic, and pre-annotated sets of marker genes from multiple sources. Mouse marker genes for the five brain cell-types, microglia, astrocytes, oligodendrocytes, neurons, and endothelial cells were obtained from Neuroexpresso (Mancarci et al. 2017) and converted to their human homologues using homologene (Mancarci, 2019). Marker genes for cellular pathways included the hallmark gene sets from the molecular signatures database (MSigDB) (Liberzon et al. 2015). The six sets of glial activation genes, microglial activation, disease-associated microglia (DAM), disease-associated astrocytes (DAA), astrocyte reactivity (MCAO and LPS), and plaque-induced genes were obtained from their respective websites and supplementary materials (Keren-Shaul et al. 2017; Habib et al. 2020; B. O. Mancarci et al. 2017; Chen et al. 2020; Zamanian et al. 2012). For each combination of brain region DEGs and marker genes, GSEA ranks the DEGs by t-statistic, which has the sign of the direction of differential expression, and takes a running cumulative tally of the overlap with the genes in the set. The maximal score is the enrichment score (ES), which is a measure of how enriched a gene set is at the top or bottom of the ranked DEG list. The DEG list is randomly shuffled and the process repeated to generate an empirical null ES distribution to calculate a P-value. To compare between each gene set tested, each ES is divided by the mean null ES to create a normalised enrichment score (NES). A positive NES means a gene set is enriched at the top of the ranked list. All results from GSEA are provided in **Supplementary Table 4**.

### Expression-weighted cell-type enrichment analysis

Expression-weighted cell-type enrichment analysis was performed using the EWCE package (Skene and Grant 2016). We performed two separate analyses using the reference single-cell RNA-seq datasets by Mathys et al (Mathys et al. 2019) and Darmanis et al (Darmanis et al. 2015). Using the reference single-cell data, cell-type specificity scores were calculated for the top 250 upregulated and downregulated genes for each brain region, ordered by t-statistic. The specificity scores of each set were then compared to the mean of the empirical null distribution from 10,000 randomly sampled gene lists. For each comparison, enrichment was expressed as the number of standard deviations from the mean. P-values were Bonferroni-corrected before applying a significance threshold of adjusted P < 0.05.

### Cell-type deconvolution

Cell-type deconvolution was performed on the voom-normalised RNA-seq data using the reference datasets by Mathys et al and Darmanis et al (Darmanis et al. 2015; Mathys et al. 2019). We first ran the dtangle package (Hunt et al. 2019) to estimate cell-type proportions in the FTLD patients and controls. After regressing out the same clinical and technical variables as in the differential expression modelling, we applied the Wilcoxon rank sum test to compare the estimated proportions of each cell-type between the patients and controls. For all comparisons, P-values were Bonferroni-corrected for multiple-testing, and significance was set at adjusted P < 0.05. Correlating the dtangle estimates from the two reference datasets, we identified strong associations across the four overlapping cell-types **(Supplementary Fig. 13)**. Using the Darmanis reference, we also ran the MuSiC algorithm, a state-of-the-art deconvolution method that accounts for cross-subject variance in gene expression (Wang et al. 2019). The estimated proportions from MuSiC and dtangle were highly correlated, although the magnitude of the estimates differed considerably (**Supplementary Fig. 12**). Deconvolution results from dtangle and Mathys et al are presented in **Supplementary Table 7**.

### Correlations with neuropathological atrophy scores

Atrophy scores for the frontal and temporal regions were manually determined by pathologists at the UCL Queen Square Brain Bank. The cortical samples were graded for both macroscopic and microscopic atrophy using the following ordinal scale: (0) absent, (1) mild, (2) moderate, and (3) severe atrophy. Macroscopic atrophy was determined from observations of gyri and sulci from the coronal slices observed during brain cutting procedures. Microscopic atrophy was determined by assessing the amount of neuronal loss on hematoxylin and eosin stained sections of the cortical brain regions. **Supplementary Table 8** lists the atrophy scores for the subset of 45 FTLD-TDP samples with RNA-seq data, and includes whether atrophy is symmetrical between both hemispheres or is less or more severe on the hemisphere used for sequencing.

We then correlated the neuropathological scores of the frontal and temporal regions with the estimated proportions of each cell-type (excitatory neurons, inhibitory neurons, endothelial cells, pericytes, astrocytes, oligodendrocytes, and microglia). For both microscopic **(Supplementary Fig. 14)** and macroscopic atrophy **(Supplementary Fig. 15)**, we used Wilcoxon rank sum tests to compare the proportions between each successive atrophy stage.

### Correlations with microglial burden scores

Microglia burden scores for a subset of the FTLD-TDP samples (**Supplementary Table 10)** were extracted from the supplementary data provided by Woollacott et al (Woollacott et al. 2020). Briefly, frontal and temporal tissue was fixed and immunohistochemically stained with antibodies against IBA1, CD68 and CR3/43 and 3,3′-Di-aminobenzidine (DAB) was used as the chromogen. Slides were scanned and digitised for analysis with ImageJ. For each of the three microglial markers, the number of positive cells were divided by the total number of DAB-positive cells in each section to construct burden scores. These scores were then correlated with the estimated microglial proportions **(Supplementary Fig. 16)**.

### Correlations with DNA methylation estimates of neuronal proportion

Genomic DNA was extracted from flash-frozen frontal cortex grey matter tissue using standard protocols. A bisulfite conversion was performed with 500 ng of genomic DNA using the EZ DNA Methylation Kit (Zymo Research). Genome-wide methylation profiling was performed using the Infinium HumanMethylationEPIC BeadChip (Illumina), as per the manufacturer’s instructions. Beta values were used to estimate the methylation levels of each CpG site using the ratio of intensities between methylated and unmethylated alleles. Beta values range from 0 to 1, representing approximately 0% to 100% methylation, respectively. Data analysis was performed using several R Bioconductor packages. Briefly, raw data (idat files) were imported into R for pre-processing and quality control using minfi (Aryee et al. 2014), ChAMP (Tian et al. 2017), and watermelon (Pidsley et al. 2013). Probes that met one or more of the following criteria were excluded from further analysis: 1) poor quality, 2) cross-reactive, 3) overlapped common genetic variants, and 4) mapped to X or Y chromosome. Samples were dropped during quality control if: 1) presenting with high failure rate, 2) the predicted sex was not matching the phenotypic sex, and 3) inappropriately clustering on multidimensional scaling analysis. Beta values were normalised with ChAMP using the Beta-Mixture Quantile (BMIQ) Normalisation method, and neuronal proportions were then estimated using the CETS package (Guintivano, Aryee, and Kaminsky 2013). For the subset of 17 FTLD-TDP samples with DNA methylation and RNA-seq data, the neuronal proportion estimates from DNA methylation **(Supplementary Table 9)** were correlated with excitatory and inhibitory neuron estimates from gene expression **(Supplementary Fig. 17)**.

### Differential Transcript Usage Analysis

Transcript expression was estimated in each sample using RSEM with the GENCODE v30 transcript reference. Lowly expressed transcripts were removed with the threshold transcript counts per million > 1 in at least 30% of all samples. Differential transcript usage (DTU) was compared between FTLD-TDP cases and controls in each brain region using satuRn (Gilis et al. 2021), a fast method for computing differential transcript usage. The same clinical and technical covariates were used as in the differential expression modelling. Pairwise comparisons between cases and controls were extracted using the limma::makeContrasts() function. We then filtered the data using a FDR threshold of 0.05, which yielded totals of 4637, 5098, and 8249 transcripts from the frontal cortex, temporal cortex, and cerebellum respectively. DTU results are presented in **Supplementary Table 3**, which includes effect sizes, P-values, and adjusted P-values for all transcripts tested.

To understand the functional profiles of the DTU genes (gDTUs), we performed a series of hypergeometric enrichment tests. We split the genes into two sets: gDTUs only, and gDTUs shared with DEGs. Using the clusterProfiler::enricher() function, we compared each set against a panel of marker genes representing the same cell-types, glial activation states, and cellular pathways as in the gene-set enrichment analyses. P-values from these enrichment tests are presented in **Supplementary Tables 5 & 6**.

### Comparisons with TDP-43 knockdown genes

Differential expression results were obtained from a TDP-43 knockdown study in IPS-derived cortical neurons (Brown et al. 2021). The results included P-values and expression log_2_ fold-changes of all genes tested, as well as Boolean values (TRUE/FALSE) indicating whether or not a gene is predicted to contain a cryptic exon. DEGs (adjusted P < 0.05) from TDP-43 knockdown were compared with DEGs from the FTLD-TDP vs Control comparison in each brain region **(Supplementary Fig. 18a)**. In addition, genes flagged as having cryptic exons were compared with DTU genes **(Supplementary Fig. 18b)**. P-values and odds ratios of all overlaps were computed using a one-sided Fisher’s exact test. Log_2_-fold changes between the FTLD-TDP and TDP-43 knockdown genes were compared using Spearman correlations **(Supplementary Fig. 18c)**.

## Supporting information

Supplementary Tables 1-14 and Supplementary Acknowledgments

## Data Availability

All raw RNA-seq data can be accessed via the NCBI GEO database (GEO GSE137810, GSE124439, GSE116622, and GSE153960). All RNA-seq data generated by the NYGC ALS Consortium are made immediately available to all members of the Consortium and with other consortia with whom we have a reciprocal sharing arrangement. To request immediate access to new and ongoing data generated by the NYGC ALS Consortium and for samples provided through the Target ALS Postmortem Core, complete a genetic data request form at.

https://www.ncbi.nlm.nih.gov/geo/query/acc.cgi?acc=GSE137810

## Code availability

All code written for this project is hosted as Rmarkdown files on Github: https://github.com/jackhump/FTLD-TDP_analysis

## Author contributions

JH and TR conceived and designed the project. RH led the main data analysis, under the supervision of JH. CB performed methylation analysis and neuronal estimation. TL provided pathological, genetic, clinical information, microglial analysis scores on the samples and performed the atrophy analyses. JH and TR oversaw all aspects of the study, with input from CB, TL, and PF. The NYGC ALS Consortium and the Target ALS Human Postmortem Tissue Core provided human tissue samples. RH and JH wrote the manuscript with input from all co-authors.

## Acknowledgements

We thank all members of the Raj lab for their feedback on the manuscript. JH and TR are funded by grants from the US National Institutes of Health (NIH NIA R56-AG055824 and NIA U01-AG068880). PF is funded by the UK MRCl (MR/M008606/1 and MR/S006508/1), the UK Motor Neurone Disease Association, Rosetrees Trust and the UCLH NIHR Biomedical Research Centre. CB is funded by the Alzheimer’s Research UK (ARUK-RF2019B-005) and the Multiple System Atrophy Trust. TL is supported by an Alzheimer’s Research UK Senior fellowship. This work was supported in part through the computational resources and staff expertise provided by Scientific Computing at the Icahn School of Medicine at Mount Sinai. Research reported in this paper was supported by the Office of Research Infrastructure of the National Institutes of Health under award number S10OD018522 and S10OD026880. All NYGC ALS Consortium activities are supported by the ALS Association (ALSA, 19-SI-459) and the Tow Foundation. The funders had no role in study design, data collection and analysis, decision to publish or preparation of the manuscript.

## Results

### Differential gene expression analyses reveal changes across multiple brain regions

We first identified general gene expression patterns in the FTLD-TDP patients by performing principal component analysis (PCA) **(Fig. 1a)**. The cerebellar samples formed a distinct cluster from the cortical samples, as expected due to cell-type differences between the cortex and cerebellum. The 2nd principal component (16.4% variance explained) indicated a clear separation between the FTD cases and controls in all three brain regions. Differentially expressed genes (DEGs) were then computed (adjusted P < 0.05) for each brain region by comparing the FTLD-TDP patients with controls, while adjusting for clinical and technical variation **(see Methods)**. We observed widespread changes in gene expression across the brain, with the largest number of unique genes observed in the frontal cortex (1711 DEGs), followed by temporal cortex (709 DEGs) and cerebellum (438 DEGs). For all the brain regions, we overlapped the FTLD-TDP DEGs **(Fig. 1b)**, and found that the frontal and temporal cortex shared the largest number of genes (FDR < 0.05 in both regions), while overlaps between cortical tissues and the cerebellum were lower. We correlated the log_2_-fold change effect sizes of each gene tested between each pair of brain regions **(Fig. 1d)**. The frontal and temporal cortex showed a strong positive correlation (R = 0.840), suggesting that these two areas have similar gene expression profiles in FTLD-TDP. In the correlations involving the cerebellum, the directionality of the expression changes were concordant, but the effect sizes were weaker (R = 0.476 with frontal cortex; R = 0.468 with temporal cortex), implicating both shared and distinct gene expression changes in the cerebellum.

**Fig. 1.**
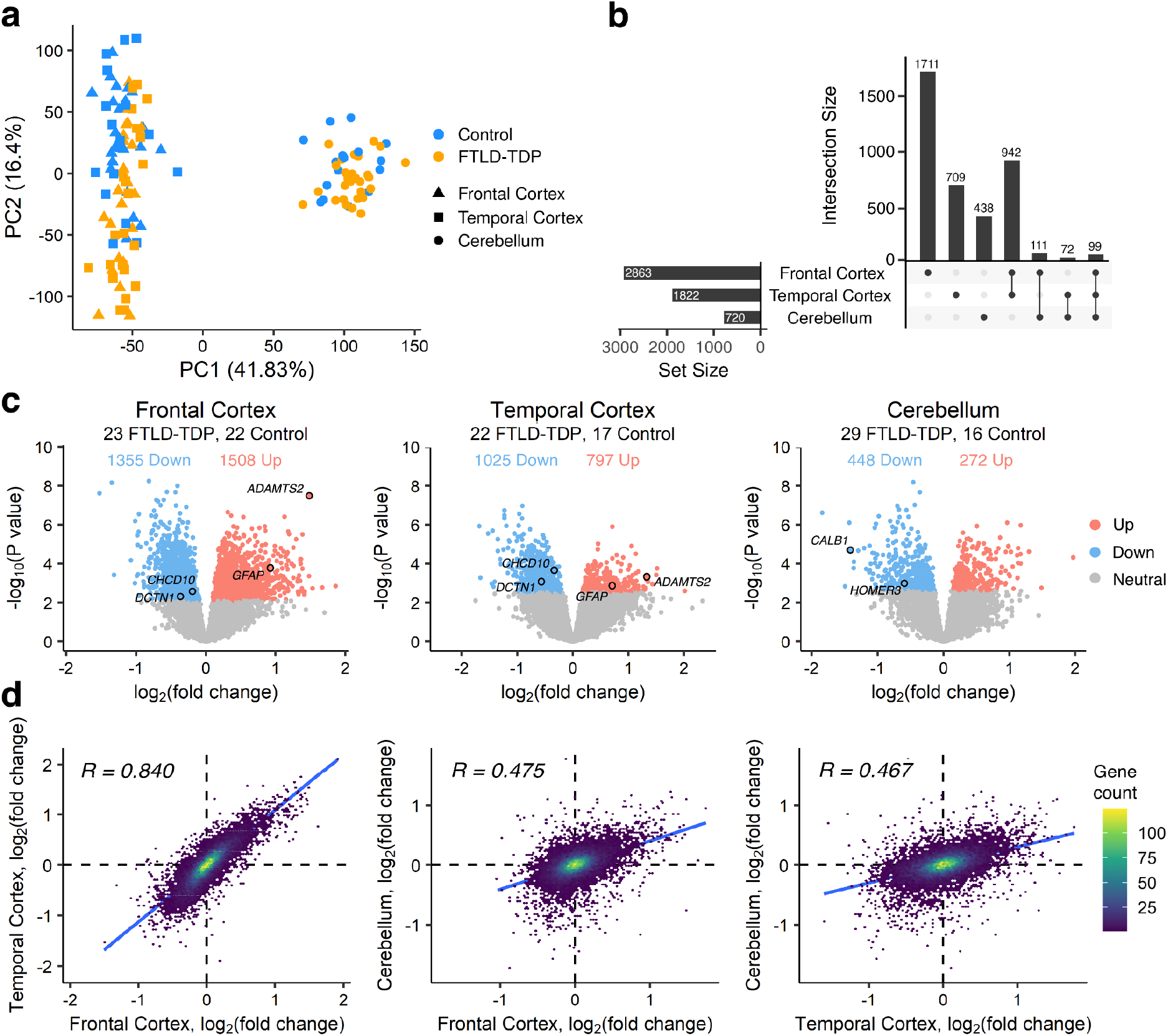
Overview of differential expression analyses. **a**. Principal component analysis of the RNA-seq expression matrix, following TMM-normalisation and covariate adjustment. Colour corresponds to disease status and shape corresponds to brain region. Distinct clusters can be seen between the cortical and cerebellar samples. **b**. Upset plot showing the number of distinct and overlapping differentially expressed genes in each brain region. **c**. Volcano plots comparing FTLD-TDP samples with controls. Red and blue dots represent genes that are upregulated and downregulated respectively (FDR adjusted P < 0.05), while gray dots are genes that are not differentially expressed. Key genes related to FTD or ALS are labelled. **d**. Scatter plots comparing log_2_-fold changes of all genes tested between each pair of brain regions. Each point is a gene, coloured by the density of overlapping points. Log_2_-fold changes are highly concordant between the frontal and temporal cortex, but less so between the cerebellum.

We identified genes that were previously implicated with FTD and/or ALS **(Fig. 1c)** including *CHCHD10, DCTN1, GFAP* and *ADAMTS2. CHCHD10* encodes a mitochondrial protein, and missense mutations in this gene have been observed in familial FTD cases and associated with impaired oxidative phosphorylation (Bannwarth et al. 2014). *DCTN1*, which encodes a motor protein involved in vesicular transport, has been shown to be downregulated in FTD and ALS (Menden et al. 2021; Kuźma-Kozakiewicz et al. 2013). *GFAP* is a marker of astrocytes previously reported to be upregulated in FTD (Hallmann et al. 2017), and the *ADAMTS2* gene encodes an enzyme responsible for collagen production (Hurskainen et al. 1999). A recent study has identified *ADAMTS2* as a marker of Von-economo neurons, a neuronal subclass vulnerable to TDP-43 pathology (Hodge et al. 2020). In the cerebellum, we highlight two downregulated genes linked to cerebellar Purkinje neurons, *CALB1* and *HOMER3*, which have been implicated in motor disorders such as Spinocerebellar ataxia and Huntington’s disease (Barski et al. 2003; Mizutani et al. 2008; Ruegsegger et al. 2016; Afshar et al. 2017).

To check for confounding effects due to post-mortem interval (PMI), we performed DGE analysis using PMI as a continuous variable. In neither control nor FTLD samples could we find any significant PMI-related genes (FDR < 0.05) in any brain region. In addition, we correlated the log_2_-fold changes between all FTLD-related and PMI-related genes **(Supplementary Fig. 4)**. Although we found positive correlations in each region, the effect sizes associated with PMI were negligible.

### C9orf72 and non-C9orf72 cases have similar gene expression profiles

We repeated DGE analysis but split the FTLD-TDP cases by *C9orf72* repeat expansion status. This allowed us to compute the gene expression changes, with respect to controls, in the 9 *C9orf72* and 21 *non-C9orf72* FTLD-TDP donors, the latter mostly consisting of sporadic disease. In the frontal cortex, most DEGs were shared between the two disease groups **(Supplementary Fig. 5a)**. Gene expression changes between the two groups were strongly concordant across the regions (frontal cortex: R = 0.89; P < 2.2e-16; temporal cortex: R = 0.90; P < 2.2e-16; cerebellum: R = 0.76; P < 2.2e-16) **(Supplementary Fig. 5b)**. Taken together, these findings are contrary to studies that have reported distinct sets of differentially expressed genes in *C9orf72* repeat expansion carriers and sporadic FTD or ALS patients (Prudencio et al. 2015; Dickson et al. 2019). Furthermore, we observed that the *C9orf72* gene itself was significantly downregulated in the *C9orf72* cases, which could be explained by hypermethylation of the *C9orf72* promoter locus (Jackson et al. 2020).

### Pathway analysis finds upregulated inflammatory response and circulatory system

Next, we performed gene set enrichment analysis (GSEA) (Subramanian et al. 2005) to examine affected cellular pathways in the FTLD-TDP patients **(Fig. 2a; Supplementary Fig. 6)**. In all three brain regions, the upregulated genes were strongly enriched for epithelial mesenchymal transition, an extracellular matrix (ECM) remodelling process, and circulatory system pathways such as angiogenesis, heme metabolism and coagulation. We also examined the expression-fold changes of matrix metalloproteinases (MMPs), which have emerged as important regulators of the ECM and circulatory system (Rempe, Hartz, and Bauer 2016). Increased expression levels of MMPs have been previously reported in various neurodegenerative diseases (Duits et al. 2015; Lu et al. 2011). Supporting a strong involvement in FTLD, we have found that most MMP genes were differentially upregulated in the cortex but not the cerebellum, and that two genes, *MMP2* and *MMP14*, were among the DEGs with the largest log_2_-fold changes **(Supplementary Fig. 7)**.

**Fig. 2.**
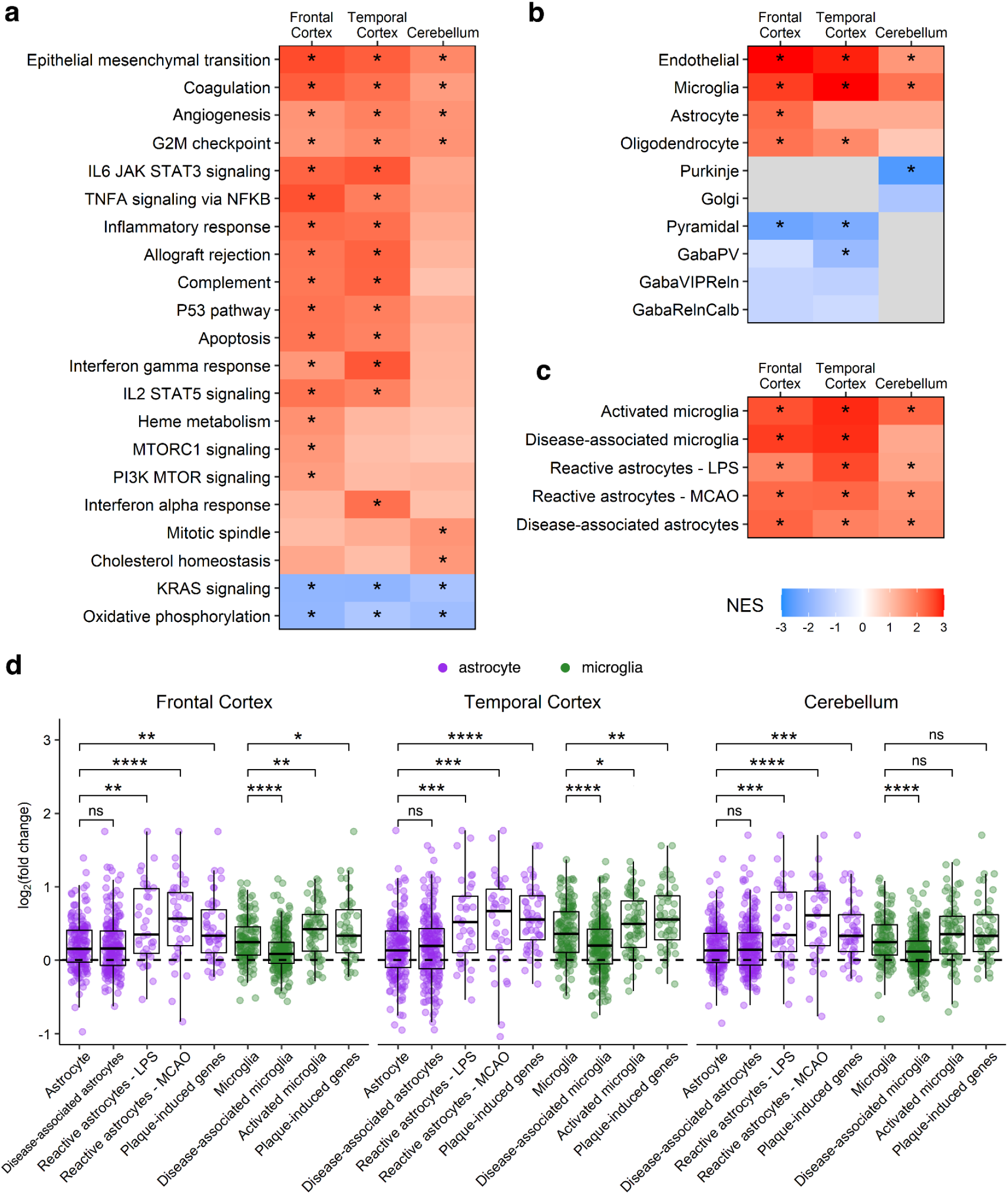
Cellular and pathway-level enrichment analyses. **a-c**. Heatmaps showing the GSEA results for cellular pathways, cell-types, and glial activation states. Coloured tiles represent the normalised-enrichment score (NES) for each term. Grey tiles were not tested. **d**. Boxplots comparing the log_2_-fold changes between the activation genes in **c** and standard marker genes in **b** for astrocytes and microglia. Comparisons were made using the Wilcoxon rank sum test. All P-values from **a-d** are Bonferroni-corrected. *** p < 1e-4; ** p < 1e-3; * p < 0.05; ns p > 0.05.

Furthermore, we observed that pathways related to immune signaling and inflammatory response were positively enriched in the cortex, but not in the cerebellum. TNF-alpha signaling via NFkB, which is believed to trigger microglia-induced neurodegeneration (Mattson and Meffert 2006), displayed the strongest enrichment among the immune signaling pathways. Accordingly, we found strong involvement of p53 pathways and apoptosis, which may act together with TDP-43 to mediate neuronal cell death (Vogt et al. 2018). Cell-proliferation pathways such as MTORC1 and PI3K/AKT/MTOR signaling were upregulated in the frontal cortex, while mitotic spindle and cholesterol homeostasis were upregulated in the cerebellum.

### Significant alterations in endothelial, glial, and neuronal gene expression

To understand how gene expression in individual brain cell-types is altered in patients with FTLD-TDP compared to controls, we performed GSEA using marker genes (Mancarci et al. 2017) for five cell types (microglia, astrocytes, oligodendrocytes, neurons, and endothelial cells) **(Fig. 2b)**. Endothelial genes were upregulated in all three brain regions and showed the strongest enrichment of the five cell-types. Enrichment for glial-cell types varied across the brain, with microglial genes upregulated in all three regions, astrocyte genes upregulated in only the frontal cortex, and oligodendrocyte genes showing increases in both the frontal and temporal cortex.

We also observed an overall reduction in neuronal gene expression. Examining the individual neuronal subtypes, Purkinje neuron expression decreased in the cerebellum, and pyramidal neuron expression decreased in the cortex. Our expression-weighted cell-type enrichment (EWCE) (Skene and Grant 2016) analysis yielded similar results **(Supplementary Fig. 8)**, showing strong loss of excitatory neuronal markers in the cortices and loss of inhibitory neuronal markers in the cortices and cerebellum. These changes could be explained by general nervous system defects caused by the disease and/or loss of neurons in the FTLD samples. A recent single-cell RNA-seq study has defined a set of marker genes for Von-economo neurons (VENs), a population of excitatory cortical neurons that are selectively reduced in TDP-43 pathology (Hodge et al. 2020). Contrary to the known vulnerability of VENs, we discovered that most of the marker genes were differentially upregulated, with the upregulated genes exhibiting larger expression fold changes than downregulated genes **(Supplementary Fig. 9)**.

To further characterise glial cell-type expression, we calculated enrichment for activated glia using a panel of immune marker genes from past transcriptomic studies (**Fig. 2c**). The gene sets, which include activated microglia, disease-associated microglia (DAM) and astrocytes (DAA), middle cerebral artery occlusion (MCAO) and lipopolysaccharide (LPS)-reactive astrocytes, and plaque-induced genes (Keren-Shaul et al. 2017; Habib et al. 2020; B. O. Mancarci et al. 2017; W.-T. Chen et al. 2020; Zamanian et al. 2012), represent signatures of microglia and astrocyte responses to a range of inflammatory stimuli. We found that the six gene sets only partially overlapped (**Supplementary Fig. 10**) and all were upregulated in the three brain regions. To understand which activation signatures disrupt glial homeostasis and are likely to play a role in inflammation, we compared the log_2_-fold changes of the activation genes with those of the standard marker genes (**Fig. 2d**). Comparing the microglial activation sets with the microglia markers in each region, we observed higher log_2_-fold changes for plaque-induced genes and activated microglia, but lower log_2_-fold changes for DAM genes. In the comparisons involving astrocytes, plaque-induced genes, and the two astrocyte reactivity sets (MCAO and LPS) exhibited higher fold changes, but DAA genes did not show any significant differences. These results point to the involvement of distinct microglia and astrocyte signatures which may play roles in the inflammatory response seen in FTLD-TDP.

### Altered compositions of endothelial cells, microglia, and neurons

Next, to estimate changes in cellular composition, we performed deconvolution analysis using a human cortical snRNA-seq dataset (Mathys et al. 2019). After adjusting for clinical and technical variation **(see Methods)**, we compared the cellular proportion estimates between the control and FTLD-TDP samples using the Wilcoxon rank sum test **(Fig. 3a)**. Endothelial cells and pericyte proportions increased in all three regions, while excitatory and inhibitory neurons were decreased in the cortex. Inhibitory neurons were also reduced in the cerebellum, consistent with the marker gene expression changes for inhibitory Purkinje neurons. Notably, proportions of microglia, astrocytes, and oligodendrocytes were increased in the cortices, but not in the cerebellum, contrary to our GSEA results with marker genes. This could be due to the fact we used cellular markers from the human cortex, which may not be fully representative of the cerebellum, or due to the use of covariate adjustment for the GSEA but not the deconvolution. Overall, the composition changes were broadly consistent with the GSEA **(Fig. 2C)** and EWCE **(Supplementary Fig. 8)** results. Using a different deconvolution tool and another reference dataset (Darmanis et al. 2015), we obtained similar results for the overlapping cell-types **(Supplementary Fig. 11-13)**.

**Fig. 3.**
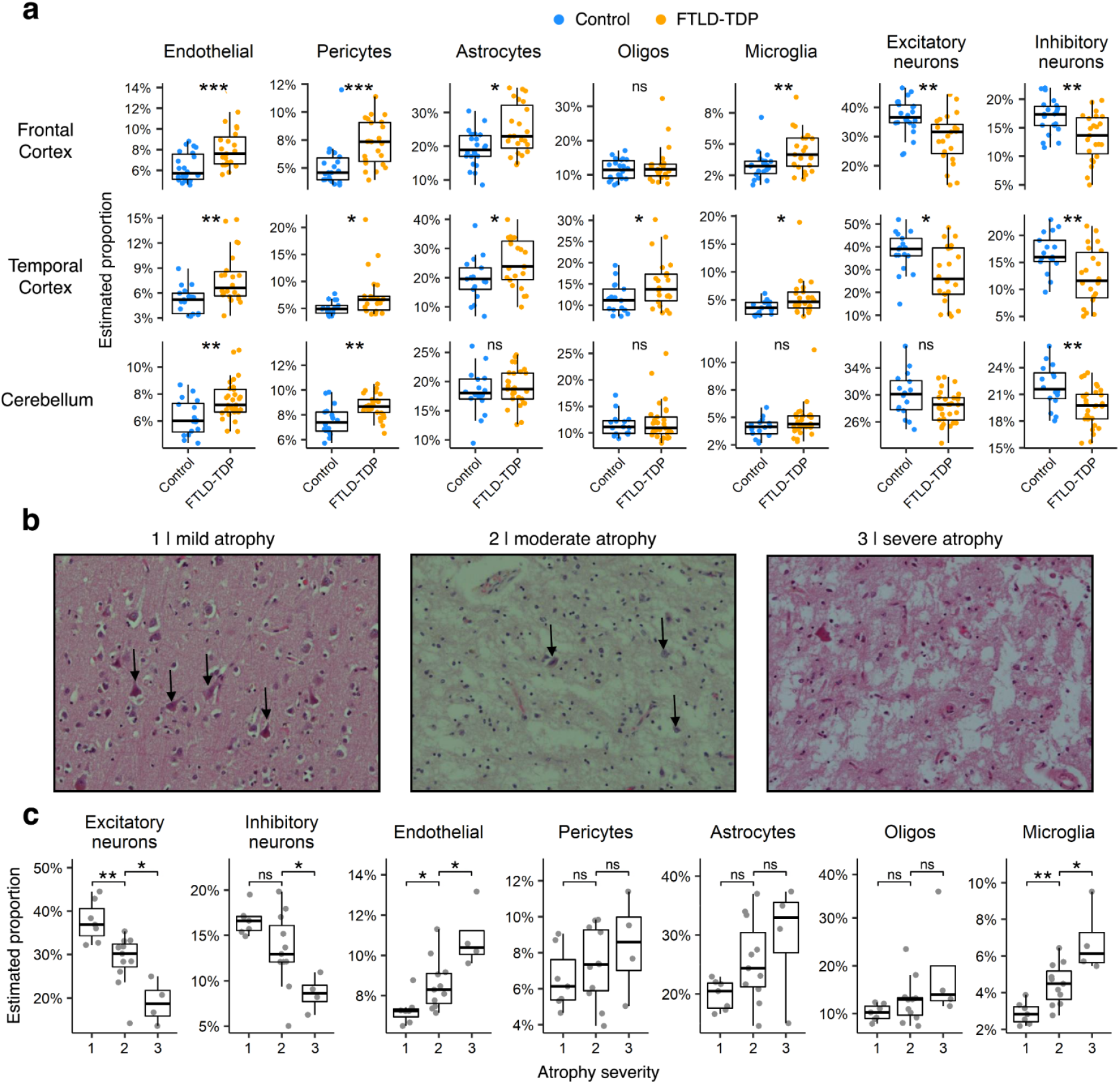
Cellular proportion changes and atrophy correlations. **a**. Comparisons of the cellular proportion estimates between control and FTLD-TDP samples. Estimates were generated with dtangle using single nuclear RNA-seq data from Mathys et al. **b**. Hematoxylin and eosin stained sections of select frontal cortex FTLD-TDP samples. From left to right, the slides depict cases with mild, moderate, and severe microscopic atrophy. Neurons are indicated by black arrows. **c**. Comparisons of the cellular proportion estimates of the frontal cortex FTLD-TDP samples between each microscopic atrophy stage. Excitatory neurons are associated with neuronal loss as the estimates show significant decreases between all atrophy stages. Inhibitory neurons show decreases between stages 2 and 3, but not 1 and 2, indicating a partial negative association. Positive associations with neuronal loss are observed for endothelial cells and microglia. Asterisks in **a** and **c** represent Bonferroni-adjusted P-values from Wilcoxon rank sum tests. *** p < 1e-4; ** p < 1e-3; * p < 0.05; ns p > 0.05.

To validate our deconvolution results, we correlated the estimated neuronal proportions for each sample with manually determined atrophy scores from a neuropathologist, determined by assessing the amount of neuronal loss in microscopic **(Fig. 3b; Supplementary Fig. 14)** or macroscopic observations of the cortical samples **(Supplementary Fig. 15)** and grouping samples into stages. In the correlations involving macroscopic atrophy, we failed to identify associations in either the frontal or temporal cortex. However, we observed significant correlations with the microscopic atrophy scores of the frontal cortex samples **(Fig. 3c)**. Both excitatory and inhibitory neuronal proportions displayed strong negative associations (**Fig. 3c)** with microscopic atrophy, thereby providing experimental confirmation of our computational predictions. Excitatory neurons were more strongly associated with the scores than inhibitory neurons, suggesting that in the frontal cortex, neuronal loss can be mainly attributed to a loss of excitatory neurons. Conversely, endothelial and microglial cells showed strong positive associations with atrophy scores. These findings may indicate a potential relationship between neuronal loss and microglial and endothelial cell changes in the frontal cortex.

Similarly to using gene expression, DNA methylation of purified cell-types can be used to deconvolve the cell-type composition of mixed samples. We used methylation calls for 17 of the FTLD-TDP frontal cortex donors to estimate neuronal proportions in those samples. However, these estimates showed no correlation with estimates derived from gene expression (R = 0.15; P = 0.56; **(Supplementary Fig. 16)**).

Additionally, we compared the estimated microglial proportions with histologically determined microglial proliferation scores on a subset of the same samples from a previous study (Woollacott et al. 2020). 14 frontal cortex and 13 temporal cortex samples were shared between the two studies. Each sample was stained for IBA1, a constitutive marker for microglia, CD68, a marker of activated phagocytic microglia, and CR3/43, which detects major compatibility complex class II molecules including HLA-DR, HLA-DP and HLA-DQ, present in activated antigen-presenting microglia. When correlating counts and combined counts of cells positive for any of the three stains, no measure significantly correlated with microglia proportion **(Supplementary Fig. 17)**.

### Differential transcript usage analysis highlights additional genes and pathways

To further characterise differences in gene expression between the FTLD-TDP patients and controls, we applied differential transcript usage (DTU) analysis. This allowed us to identify expression changes, at the level of transcripts, caused by alterations in polyadenylation, promoter usage, and alternative splicing. Adjusting for clinical and technical variation **(see Methods)**, the analysis revealed 2630, 2891, and 4045 genes with differential transcript usage (gDTU) in the frontal cortex, temporal cortex, and cerebellum respectively **(Fig. 4a)**. In addition, most gDTUs did not overlap with the DEGs in any brain region **(Fig. 4a)**.

**Fig. 4.**
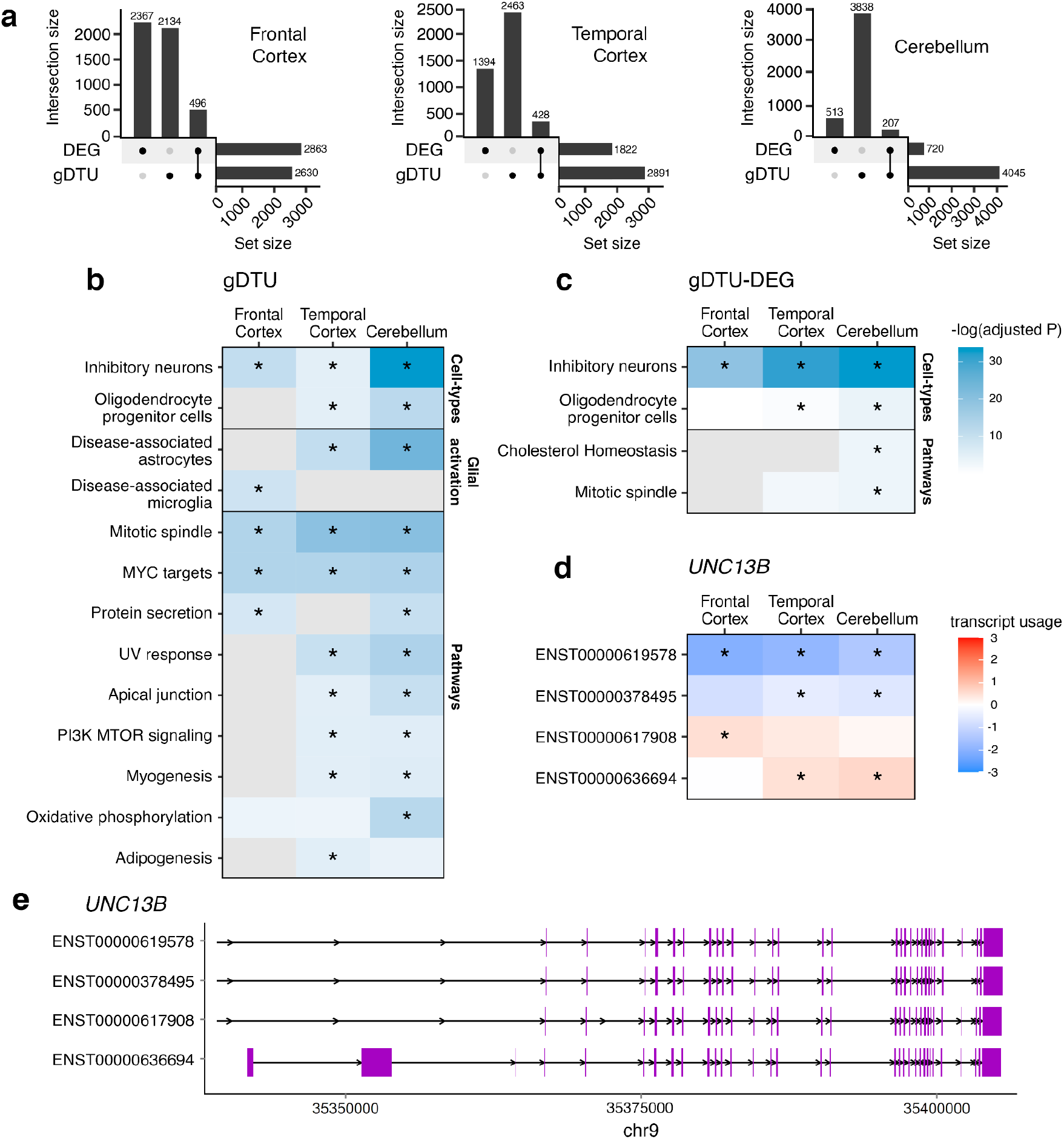
Differential transcript usage analysis. **a**. Upset plots showing overlaps between the gDTUs (adjusted P < 0.05) and DEGs (adjusted P < 0.05) for each brain region. **b-c**. Cell-types, glial activation states, and pathways enriched in gDTUs **(b)** and gDTUs + DEGs **(c)**. Coloured tiles represent the negative logarithm of the adjusted P-value for each term. Grey tiles were not tested. **d**. Transcript usage of the four *UNC13B* isoforms with significant effect sizes in at least one brain region. **e**. Alignment chart of the *UNC13B* isoforms in **d**, showing the region with greatest variation in transcript structure. Purple rectangles correspond to protein-coding exons. Asterisks in **b-d** refer to P-values that meet the FDR threshold of 0.05. *** p < 1e-4; ** p < 1e-3; * p < 0.05; ns p > 0.05.

To understand the functional consequences of TDP-43 pathology, we performed a series of enrichment tests on the gDTUs. We split the genes into two sets: gDTUs only, and gDTUs shared with DEGs. We then compared each set against a panel of marker genes representing the same cell-types, glial activation states, and cellular pathways as before **(Fig. 4b-c)**. gDTUs, either alone or in combination with DEGs, showed significant enrichment (adjusted P < 0.05) for inhibitory neurons and oligodendrocyte progenitor cells in at least one brain region. Inputting only gDTUs, we found a strong presence of DAM genes in the frontal cortex, and DAA genes in the temporal cortex and cerebellum. We also identified several pathways in certain regions that were overlooked by our previous enrichment analysis using DEGs, such as mitotic spindle and apical junction, two pathways used by non-neuronal cell types. We show the transcript-level expression changes of *UNC13B*, a synaptic gene which may have important functional relevance in FTLD-TDP **(Fig. 4d)**. Brown and colleagues have previously reported a loss of *UNC13B* expression due to splicing changes caused by TDP-43 knockdown in neuronal cell lines (Brown et al. 2021). Elaborating on this finding, we observed that *UNC13B* was downregulated in the cortices, and that four of the six *UNC13B* transcripts were altered in at least one brain region. The four transcripts differ by cassette exon inclusion, 3’UTR length and alternative promoter usage. Usage of ENST00000619578 decreased across all three brain regions, and the effect sizes associated with this isoform were among the largest in the transcriptome. Usage for ENST00000378495 also decreased within the temporal cortex and cerebellum, while that of ENST000061708 and ENST0000036694 increased.

The finding of DTU of *UNC13B* led us to consider whether changes in gene expression and splicing directly related to TDP-43 pathology might be observed in our bulk brain samples. TDP-43 nuclear loss pathology should be present in both frontal and temporal cortex but is not generally observed in the cerebellum (Neumann et al. 2006). Recently, RNA-seq on human iPSC-derived neurons identified differentially expressed genes resulting from TDP-43 knockdown, as well as genes with possible cryptic exon (CE) inclusions (Brown et al. 2021). We compared the DEGs from our FTLD-TDP vs control comparison with those found in the study by Brown et al **(Supplementary Fig. 18a)**. These TDP-43 knockdown genes overlapped with at least 25% of the FTLD-TDP DEGs found in each tissue (frontal cortex: OR = 1.23; P = 1.65e-5; temporal cortex: OR = 1.42; P = 1.12e-9; cerebellum: OR = 1.64; P = 1.02e-8). Furthermore, we overlapped our gDTUs with genes with possible CE inclusions and identified 52, 58, and 72 genes in the frontal cortex, temporal cortex, and cerebellum respectively **(Supplementary Fig. 18b)**. By correlating the expression-fold changes between the two gene sets **(Supplementary Fig. 18c)**, we also found weak, but nonetheless significant associations (P < 0.05) in all three regions (frontal cortex: R = 0.14; P < 2.2e-16; temporal cortex: R = 0.13; P < 2.2e-16; cerebellum: R = 0.058, P = 3.2e-11).

## Discussion

In this study, we assembled a large cohort of RNA-seq from post-mortem FTLD-TDP brains to understand the cellular mechanisms underlying FTLD and TDP-43 pathology. We provide a detailed landscape of gene expression alterations from multiple brain regions, highlighting the roles of specific glial cell-types, the vulnerability of excitatory neurons, and a strong involvement of the cerebellum in the neurodegenerative process of FTLD. Our data represents a transcriptomic resource to help accelerate future research towards a better understanding of the pathogenic mechanisms in FTD.

By profiling transcriptomic changes in FTLD brains using multiple techniques (GSEA, EWCE, deconvolution), we identified robust shifts in cell-type abundance and expression across both the cortex and cerebellum. Notably, we show that microglia, astrocytes, and oligodendrocytes, cell-types believed to play key roles in the neurodegenerative process of FTLD, were prominently upregulated in the frontal and temporal cortex. We found that genes associated with some but not all glial activation sets exhibited stronger expression fold-changes than standard marker genes, suggesting a possible shift towards activated glia in FTLD brains. Furthermore, in line with the known consequences of glial activation (McCauley and Baloh 2019), we have identified enrichment in neuroinflammation and apoptosis pathways in the cortical regions. However, we are aware that these interpretations are limited by our use of bulk-tissue sections. A more thorough analysis of glial cell-type changes would involve quantifying the relative cellular proportions between intrinsic cell-states at the level of single cells. Moreover, we were unable to assess whether the observed changes in microglia and astrocyte gene expression are at least partly driven by peripheral monocytes, which are known to infiltrate through the blood brain barrier (da Fonseca et al. 2014).

Our enrichment analyses have detected widespread upregulation of endothelial cells and angiogenesis pathways, suggesting increased blood vessel abundance and growth in FTLD brains. It is generally not known how or if the circulatory system is involved in FTLD pathogenesis, although vascular abnormalities have been found in postmortem brains with TDP-43 pathology (Ek Olofsson and Englund 2019). In all brain regions, we also detected strong enrichment for extracellular matrix (ECM) remodelling, a cellular process that is poorly understood in FTLD. One of the few lines of evidence supporting ECM dysregulation in FTLD comes from a transcriptomic study done on individual genetic subgroups - *C9orf72, GRN*, and *MAPT* (Menden et al. 2021). In all three groups, Menden et al detected increased expression of key ECM genes, including matrix metalloproteinases (MMPs). In other neurodegenerative diseases, MMP pathways have been shown to increase production of growth factors that promote blood vessel development, providing a potential causal link between the observed enrichment of endothelial cells (Rempe, Hartz, and Bauer 2016). Given this relationship, we think the ECM and circulatory system represent important subjects to study for future FTD research.

From our deconvolution analysis, we have identified robust decreases in excitatory and inhibitory neuron proportions in the cortical regions. These findings fall in line with initial studies on postmortem FTLD brains that have found deficits to glutamatergic and GABAergic systems (Ferrer 1999). Recently, evidence from multiple studies has accumulated in support of the selective vulnerability of excitatory neurons to TDP-43 pathology (Benussi et al. 2019; Zhang et al. 2020; Santillo, Nilsson, and Englund 2013; Nana et al. 2019). This led us to examine the expression changes linked to Von Economo neurons (VENs), a group of selectively targeted excitatory neurons, using marker genes from the recent study by Hodge et. al (Hodge et al. 2020). However, we found that the marker genes were mainly upregulated. Given that we used bulk-tissue sections, our dataset is likely affected by the relatively low proportion of VEN neurons within each sample. In support of the vulnerability of excitatory neurons in FTLD-TDP, our correlations with neuropathological atrophy show stronger associations for excitatory neurons than inhibitory neurons. However, we note that significant associations were only observed for the frontal cortex.

We identified a decrease in inhibitory neurons in the cerebellum that agrees with recent reports of Purkinje neuron loss in FTD-ALS mouse models (Chew et al. 2015), despite lacking a cerebellum-specific single cell RNA-seq panel. We also detected a reduction in expression of known Purkinje neuron marker genes in our enrichment analyses. These findings add to the growing body of evidence supporting the involvement of the cerebellum in FTLD. Imaging studies have shown that the cerebellum may be selectively targeted in sporadic FTLD-TDP and in cases caused by *C9orf72* mutations (Tan et al. 2014; Chen et al. 2019). In *C9orf72* cases, accumulation of *C9orf72* dipeptide repeat protein, rather than TDP-43, may be responsible for cerebellar neuronal loss (Yousef et al. 2017; Gendron et al. 2015), although a direct causal link has not yet been established. Interestingly, by comparing FTLD-TDP cases with and without *C9orf72* mutations, we found concordant changes in gene expression in the cerebellum. Potentially, this could suggest that mechanisms driving cerebellar neuronal loss are shared between *C9orf72* and sporadic cases.

We did not observe significant correlations with atrophy scores from the temporal cortex. Nor did we observe concordance between neuronal proportions estimated from DNA methylation, nor between estimated microglia proportions and histological counts of microglia. These failed associations are presumably partly due to sample size as well as the difficulties of accurate deconvolution of brain samples (Patrick et al. 2020). For the histological metrics of microglia counts and neuronal loss, there is the additional confounder of asymmetric changes between the hemispheres, as the deconvolution estimates and neuropathological observations were determined from opposite hemispheres of the same donor. To resolve this in future, it would be useful to fix and freeze adjacent sections of the same hemisphere.

Applying DTU analysis has allowed us to detect changes in transcript usage in FTLD-TDP affecting a largely distinct set of genes from the DGE analysis. These genes were less enriched in specific cell-types, which suggests they may reflect cell-intrinsic changes, rather than simply cellular composition changes exhibited by the DEGs. For example, the largest number of genes with DTU was observed for the cerebellum, in which we previously found the lowest number of DEGs and the weakest cell-type enrichments. Multiple studies have demonstrated that reductions in nuclear TDP-43 alter the expression and splicing of its target mRNAs, many of which regulate neuronal function (Polymenidou et al. 2011; Tollervey et al. 2011; Rot et al. 2017). In support of this, we observed significant overlaps between DTU genes and genes known to be altered by TDP-43 loss. However, as widespread DTU has been observed in multiple neurodegenerative diseases, including Alzheimer’s (Marques-Coelho et al., 2020) and Parkinson’s (Dick et al. 2020), as well as similar enrichment with DTU genes found in the cerebellum, we are wary of linking the majority of transcript changes to TDP-43 pathology. For example, the DTU study of Alzheimer’s brains also observed DTU in genes and pathways related to immune activation and synaptic transmission (Marques-Coelho et al., 2020). The DTU genes may therefore reflect a general consequence of neurodegenerative disease rather than TDP-43 pathology. Another example of this is the observation of DTU in both *UNC13A* and *UNC13B*, two synaptic genes whose splicing is altered in TDP-43 loss-of-function models (Ma et al. 2021; Brown et al. 2021). For *UNC13B*, the prominent DTU signals observed in the frontal and temporal cortex would suggest that this gene is affected by loss of TDP-43. However, given that DTU signals were also observed in the cerebellum, a region that lacks TDP-43 pathology, we think some other factor, in addition to TDP-43 pathology, may converge on these critical neuronal genes.

A major limitation of this study is the sample size of the FTLD-TDP group. We lacked a sufficient number of donors to analyze differences between individual genetic or TDP-43 pathology subtypes. A larger cohort would allow us to identify cell-types and pathways that are affected between subgroups of FTLD, and grant us statistical power to perform additional analyses such as gene co-expression networks. Hopefully, in future studies we will be able to meta-analyse across FTLD cohorts.

To conclude, we highlight potential consequences of FTLD-TDP, including vascular dysfunction, RNA missplicing, and glial activation. We emphasize the vulnerability of excitatory neurons, and a strong involvement of the cerebellum in the FTLD neurodegenerative process. We hope our data stimulates further research that will lead toward a better understanding of the relevant disease mechanisms in FTLD.

## References

Afshar, Pegah, Niloufar Ashtari, Xiaodan Jiao, Maryam Rahimi-Balaei, Xiaosha Zhang, Behzad Yaganeh, Marc R. Del Bigio, Jiming Kong, and Hassan Marzban. 2017. “Overexpression of Human SOD1 Leads to Discrete Defects in the Cerebellar Architecture in the Mouse.” Frontiers in Neuroanatomy 11 (March): 22.

Aryee, Martin J., Andrew E. Jaffe, Hector Corrada-Bravo, Christine Ladd-Acosta, Andrew P. Feinberg, Kasper D. Hansen, and Rafael A. Irizarry. 2014. “Minfi: A Flexible and Comprehensive Bioconductor Package for the Analysis of Infinium DNA Methylation Microarrays.” Bioinformatics 30 (10): 1363–69.

Baker, Matt, Ian R. Mackenzie, Stuart M. Pickering-Brown, Jennifer Gass, Rosa Rademakers, Caroline Lindholm, Julie Snowden, et al. 2006. “Mutations in Progranulin Cause Tau-Negative Frontotemporal Dementia Linked to Chromosome 17.” Nature 442 (7105): 916–19.

Bannwarth, Sylvie, Samira Ait-El-Mkadem, Annabelle Chaussenot, Emmanuelle C. Genin, Sandra Lacas-Gervais, Konstantina Fragaki, Laetitia Berg-Alonso, et al. 2014. “A Mitochondrial Origin for Frontotemporal Dementia and Amyotrophic Lateral Sclerosis through CHCHD10 Involvement.” Brain: A Journal of Neurology 137 (Pt 8): 2329–45.

Barski, Jaroslaw J., Jana Hartmann, Christine R. Rose, Freek Hoebeek, Karin Mörl, Michael Noll-Hussong, Chris I. De Zeeuw, Arthur Konnerth, and Michael Meyer. 2003. “Calbindin in Cerebellar Purkinje Cells Is a Critical Determinant of the Precision of Motor Coordination.” The Journal of Neuroscience: The Official Journal of the Society for Neuroscience 23 (8): 3469–77.

Benussi, Alberto, Antonella Alberici, Emanuele Buratti, Roberta Ghidoni, Fabrizio Gardoni, Monica Di Luca, Alessandro Padovani, and Barbara Borroni. 2019. “Toward a Glutamate Hypothesis of Frontotemporal Dementia.” Frontiers in Neuroscience 13 (March): 304.

Bersano, Anna, Roberto Del Bo, Costanza Lamperti, Serena Ghezzi, Gigliola Fagiolari, Francesco Fortunato, Elena Ballabio, et al. 2009. “Inclusion Body Myopathy and Frontotemporal Dementia Caused by a Novel VCP Mutation.” Neurobiology of Aging 30 (5): 752–58.

Borroni, B., C. Bonvicini, A. Alberici, E. Buratti, C. Agosti, S. Archetti, A. Papetti, et al. 2009. “Mutation within TARDBP Leads to Frontotemporal Dementia without Motor Neuron Disease.” Human Mutation 30 (11): E974–83.

Brown, Anna-Leigh, Oscar G. Wilkins, Matthew J. Keuss, Sarah E. Hill, Matteo Zanovello, Weaverly Colleen Lee, Flora C. Y. Lee, et al. 2021. “Common ALS/FTD Risk Variants in UNC13A Exacerbate Its Cryptic Splicing and Loss upon TDP-43 Mislocalization.” bioRxiv. https://doi.org/10.1101/2021.04.02.438170.

Chen, Wei-Ting, Ashley Lu, Katleen Craessaerts, Benjamin Pavie, Carlo Sala Frigerio, Nikky Corthout, Xiaoyan Qian, et al. 2020. “Spatial Transcriptomics and In Situ Sequencing to Study Alzheimer’s Disease.” Cell 182 (4): 976–91.e19.

Chen, Yu, Fiona Kumfor, Ramon Landin-Romero, Muireann Irish, and Olivier Piguet. 2019. “The Cerebellum in Frontotemporal Dementia: A Meta-Analysis of Neuroimaging Studies.” Neuropsychology Review 29 (4): 450–64.

Chew, Jeannie, Tania F. Gendron, Mercedes Prudencio, Hiroki Sasaguri, Yong-Jie Zhang, Monica Castanedes-Casey, Chris W. Lee, et al. 2015. “Neurodegeneration. C9ORF72 Repeat Expansions in Mice Cause TDP-43 Pathology, Neuronal Loss, and Behavioral Deficits.” Science 348 (6239): 1151–54.

Cruts, Marc, Ilse Gijselinck, Julie van der Zee, Sebastiaan Engelborghs, Hans Wils, Daniel Pirici, Rosa Rademakers, et al. 2006. “Null Mutations in Progranulin Cause Ubiquitin-Positive Frontotemporal Dementia Linked to Chromosome 17q21.” Nature 442 (7105): 920–24.

Darmanis, Spyros, Steven A. Sloan, Ye Zhang, Martin Enge, Christine Caneda, Lawrence M. Shuer, Melanie G. Hayden Gephart, Ben A. Barres, and Stephen R. Quake. 2015. “A Survey of Human Brain Transcriptome Diversity at the Single Cell Level.” Proceedings of the National Academy of Sciences of the United States of America 112 (23): 7285–90.

DeJesus-Hernandez, Mariely, Ian R. Mackenzie, Bradley F. Boeve, Adam L. Boxer, Matt Baker, Nicola J. Rutherford, Alexandra M. Nicholson, et al. 2011. “Expanded GGGGCC Hexanucleotide Repeat in Noncoding Region of C9ORF72 Causes Chromosome 9p-Linked FTD and ALS.” Neuron 72 (2): 245–56.

Dick, Fiona, Gonzalo S. Nido, Guido Werner Alves, Ole-Bjørn Tysnes, Gry Hilde Nilsen, Christian Dölle, and Charalampos Tzoulis. 2020. “Differential Transcript Usage in the Parkinson’s Disease Brain.” PLoS Genetics 16 (11): e1009182.

Dickson, Dennis W., Matthew C. Baker, Jazmyne L. Jackson, Mariely DeJesus-Hernandez, Nicole A. Finch, Shulan Tian, Michael G. Heckman, et al. 2019. “Extensive Transcriptomic Study Emphasizes Importance of Vesicular Transport in C9orf72 Expansion Carriers.” Acta Neuropathologica Communications 7 (1): 150.

Duits, Flora H., Mar Hernandez-Guillamon, Joan Montaner, Jereon D. C. Goos, Alex Montañola, Mike P. Wattjes, Frederik Barkhof, Philip Scheltens, Charlotte E. Teunissen, and Wiesje M. van der Flier. 2015. “Matrix Metalloproteinases in Alzheimer’s Disease and Concurrent Cerebral Microbleeds.” Journal of Alzheimer’s Disease: JAD 48 (3): 711–20.

Ek Olofsson, H., and E. Englund. 2019. “A Cortical Microvascular Structure in Vascular Dementia, Alzheimer’s Disease, Frontotemporal Lobar Degeneration and Nondemented Controls: A Sign of Angiogenesis due to Brain Ischaemia?” Neuropathology and Applied Neurobiology 45 (6): 557–69.

Ferrer, I. 1999. “Neurons and Their Dendrites in Frontotemporal Dementia.” Dementia and Geriatric Cognitive Disorders 10 Suppl 1: 55–60.

Fonseca, Anna Carolina Carvalho da, Diana Matias, Celina Garcia, Rackele Amaral, Luiz Henrique Geraldo, Catarina Freitas, and Flavia Regina Souza Lima. 2014. “The Impact of Microglial Activation on Blood-Brain Barrier in Brain Diseases.” Frontiers in Cellular Neuroscience 8 (November): 362.

Fromer, Menachem, Panos Roussos, Solveig K. Sieberts, Jessica S. Johnson, David H. Kavanagh, Thanneer M. Perumal, Douglas M. Ruderfer, et al. 2016. “Gene Expression Elucidates Functional Impact of Polygenic Risk for Schizophrenia.” Nature Neuroscience 19 (11): 1442–53.

Gami-Patel, P., I. van Dijken, J. C. van Swieten, Y. A. L. Pijnenburg, Netherlands Brain Bank, A. J. M. Rozemuller, J. J. M. Hoozemans, and A. A. Dijkstra. 2019. “Von Economo Neurons Are Part of a Larger Neuronal Population That Are Selectively Vulnerable in C9orf72 Frontotemporal Dementia.” Neuropathology and Applied Neurobiology 45 (7): 671–80.

Gendron, Tania F., Marka van Blitterswijk, Kevin F. Bieniek, Lillian M. Daughrity, Jie Jiang, Beth K. Rush, Otto Pedraza, et al. 2015. “Cerebellar c9RAN Proteins Associate with Clinical and Neuropathological Characteristics of C9ORF72 Repeat Expansion Carriers.” Acta Neuropathologica 130 (4): 559–73.

Gijselinck, Ilse, Sara Van Mossevelde, Julie van der Zee, Anne Sieben, Stéphanie Philtjens, Bavo Heeman, Sebastiaan Engelborghs, et al. 2015. “Loss of TBK1 Is a Frequent Cause of Frontotemporal Dementia in a Belgian Cohort.” Neurology 85 (24): 2116–25.

Gilis, Jeroen, Kristoffer Vitting-Seerup, Koen Van den Berge, and Lieven Clement. 2021. “satuRn: Scalable Analysis of Differential Transcript Usage for Bulk and Single-Cell RNA-Sequencing Applications.” bioRxiv. https://doi.org/10.1101/2021.01.14.426636.

Glass, Christopher K., Kaoru Saijo, Beate Winner, Maria Carolina Marchetto, and Fred H. Gage. 2010. “Mechanisms Underlying Inflammation in Neurodegeneration.” Cell 140 (6): 918–34.

Goldman, J. S., J. M. Farmer, E. M. Wood, J. K. Johnson, A. Boxer, J. Neuhaus, C. Lomen-Hoerth, et al. 2005. “Comparison of Family Histories in FTLD Subtypes and Related Tauopathies.” Neurology 65 (11): 1817–19.

GTEx Consortium. 2020. “The GTEx Consortium Atlas of Genetic Regulatory Effects across Human Tissues.” Science 369 (6509): 1318–30.

Guintivano, Jerry, Martin J. Aryee, and Zachary A. Kaminsky. 2013. “A Cell Epigenotype Specific Model for the Correction of Brain Cellular Heterogeneity Bias and Its Application to Age, Brain Region and Major Depression.” Epigenetics: Official Journal of the DNA Methylation Society 8 (3): 290–302.

Habib, Naomi, Cristin McCabe, Sedi Medina, Miriam Varshavsky, Daniel Kitsberg, Raz Dvir-Szternfeld, Gilad Green, et al. 2020. “Disease-Associated Astrocytes in Alzheimer’s Disease and Aging.” Nature Neuroscience 23 (6): 701–6.

Hallmann, Anna-Lena, Marcos J. Araúzo-Bravo, Lampros Mavrommatis, Marc Ehrlich, Albrecht Röpke, Johannes Brockhaus, Markus Missler, et al. 2017. “Astrocyte Pathology in a Human Neural Stem Cell Model of Frontotemporal Dementia Caused by Mutant TAU Protein.” Scientific Reports 7 (March): 42991.

Hodge, Rebecca D., Jeremy A. Miller, Mark Novotny, Brian E. Kalmbach, Jonathan T. Ting, Trygve E. Bakken, Brian D. Aevermann, et al. 2020. “Transcriptomic Evidence That von Economo Neurons Are Regionally Specialized Extratelencephalic-Projecting Excitatory Neurons.” Nature Communications 11 (1): 1172.

Hoffman, Gabriel E., and Eric E. Schadt. 2016. “variancePartition: Interpreting Drivers of Variation in Complex Gene Expression Studies.” BMC Bioinformatics 17 (1): 483.

Honda, Daiyu, Shinsuke Ishigaki, Yohei Iguchi, Yusuke Fujioka, Tsuyoshi Udagawa, Akio Masuda, Kinji Ohno, Masahisa Katsuno, and Gen Sobue. 2013. “The ALS/FTLD-Related RNA-Binding Proteins TDP-43 and FUS Have Common Downstream RNA Targets in Cortical Neurons.” FEBS Open Bio 4 (November): 1–10.

Hughes, Laura E., Timothy Rittman, Trevor W. Robbins, and James B. Rowe. 2018. “Reorganization of Cortical Oscillatory Dynamics Underlying Disinhibition in Frontotemporal Dementia.” Brain: A Journal of Neurology 141 (8): 2486–99.

Hunt, Gregory J., Saskia Freytag, Melanie Bahlo, and Johann A. Gagnon-Bartsch. 2019. “Dtangle: Accurate and Robust Cell Type Deconvolution.” Bioinformatics 35 (12): 2093–99.

Hurskainen, Tiina L., Satoshi Hirohata, Michael F. Seldin, and Suneel S. Apte. 1999. “ADAM-TS5, ADAM-TS6, and ADAM-TS7, Novel Members of a New Family of Zinc Metalloproteases: GENERAL FEATURES AND GENOMIC DISTRIBUTION OF THE ADAM-TS FAMILY * 210.” The Journal of Biological Chemistry 274 (36): 25555–63.

Jackson, Jazmyne L., Nicole A. Finch, Matthew C. Baker, Jennifer M. Kachergus, Mariely DeJesus-Hernandez, Kimberly Pereira, Elizabeth Christopher, et al. 2020. “Elevated Methylation Levels, Reduced Expression Levels, and Frequent Contractions in a Clinical Cohort of C9orf72 Expansion Carriers.” Molecular Neurodegeneration 15 (1): 7.

Jung, Yeon-Joo, and Won-Suk Chung. 2018. “Phagocytic Roles of Glial Cells in Healthy and Diseased Brains.” Biomolecules & Therapeutics 26 (4): 350–57.

Keren-Shaul, Hadas, Amit Spinrad, Assaf Weiner, Orit Matcovitch-Natan, Raz Dvir-Szternfeld, Tyler K. Ulland, Eyal David, et al. 2017. “A Unique Microglia Type Associated with Restricting Development of Alzheimer’s Disease.” Cell 169 (7): 1276–90.e17.

Klim, Joseph R., Luis A. Williams, Francesco Limone, Irune Guerra San Juan, Brandi N. Davis-Dusenbery, Daniel A. Mordes, Aaron Burberry, et al. 2019. “ALS-Implicated Protein TDP-43 Sustains Levels of STMN2, a Mediator of Motor Neuron Growth and Repair.” Nature Neuroscience 22 (2): 167–79.

Kuźma-Kozakiewicz, Magdalena, Agnieszka Chudy, Beata Kaźmierczak, Dorota Dziewulska, Ewa Usarek, and Anna Barańczyk-Kuźma. 2013. “Dynactin Deficiency in the CNS of Humans with Sporadic ALS and Mice with Genetically Determined Motor Neuron Degeneration.” Neurochemical Research, September. https://doi.org/10.1007/s11064-013-1160-7.

Law, Charity W., Yunshun Chen, Wei Shi, and Gordon K. Smyth. 2014. “Voom: Precision Weights Unlock Linear Model Analysis Tools for RNA-Seq Read Counts.” Genome Biology 15 (2): R29.

Le Ber, Isabelle, Agnès Camuzat, Rita Guerreiro, Kawtar Bouya-Ahmed, Jose Bras, Gael Nicolas, Audrey Gabelle, et al. 2013. “SQSTM1 Mutations in French Patients with Frontotemporal Dementia or Frontotemporal Dementia with Amyotrophic Lateral Sclerosis.” JAMA Neurology 70 (11): 1403–10.

Liberzon, Arthur, Chet Birger, Helga Thorvaldsdóttir, Mahmoud Ghandi, Jill P. Mesirov, and Pablo Tamayo. 2015. “The Molecular Signatures Database (MSigDB) Hallmark Gene Set Collection.” Cell Systems 1 (6): 417–25.

Liddelow, Shane A., Kevin A. Guttenplan, Laura E. Clarke, Frederick C. Bennett, Christopher J. Bohlen, Lucas Schirmer, Mariko L. Bennett, et al. 2017. “Neurotoxic Reactive Astrocytes Are Induced by Activated Microglia.” Nature 541 (7638): 481–87.

Ling, Shuo-Chien, Magdalini Polymenidou, and Don W. Cleveland. 2013. “Converging Mechanisms in ALS and FTD: Disrupted RNA and Protein Homeostasis.” Neuron 79 (3): 416–38.

Lu, Pengfei, Ken Takai, Valerie M. Weaver, and Zena Werb. 2011. “Extracellular Matrix Degradation and Remodeling in Development and Disease.” Cold Spring Harbor Perspectives in Biology 3 (12). https://doi.org/10.1101/cshperspect.a005058.

Mancarci, B. Ogan, Lilah Toker, Shreejoy J. Tripathy, Brenna Li, Brad Rocco, Etienne Sibille, and Paul Pavlidis. 2017. “Cross-Laboratory Analysis of Brain Cell Type Transcriptomes with Applications to Interpretation of Bulk Tissue Data.” eNeuro 4 (6). https://doi.org/10.1523/ENEURO.0212-17.2017.

Mancarci, Ogan. 2019. “Quick Access to Homologene and Gene Annotation Updates [R Package Homologene Version 1.4.68.19.3.27],” March. https://cran.r-project.org/web/packages/homologene/index.html.

Marques-Coelho, Diego, Lukas Iohan da Cruz Carvalho, Ana Raquel Melo de Farias, Jean-Charles Lambert, Marcos Romualdo Costa, and Neuroceb Brain Bank. n.d. “Differential Transcript Usage Unravels Gene Expression Alterations in Alzheimer’s Disease Human Brains.” https://doi.org/10.1101/2020.03.19.20038703.

Mathys, Hansruedi, Jose Davila-Velderrain, Zhuyu Peng, Fan Gao, Shahin Mohammadi, Jennie Z. Young, Madhvi Menon, et al. 2019. “Single-Cell Transcriptomic Analysis of Alzheimer’s Disease.” Nature 570 (7761): 332–37.

Mattson, M. P., and M. K. Meffert. 2006. “Roles for NF-κB in Nerve Cell Survival, Plasticity, and Disease.” Cell Death and Differentiation 13 (5): 852–60.

McCauley, Madelyn E., and Robert H. Baloh. 2019. “Inflammation in ALS/FTD Pathogenesis.” Acta Neuropathologica 137 (5): 715–30.

Melamed, Ze’ev, Jone López-Erauskin, Michael W. Baughn, Ouyang Zhang, Kevin Drenner, Ying Sun, Fernande Freyermuth, et al. 2019. “Premature Polyadenylation-Mediated Loss of Stathmin-2 Is a Hallmark of TDP-43-Dependent Neurodegeneration.” Nature Neuroscience 22 (2): 180–90.

Menden, K., M. Francescatto, T. Niyma, and C. Blauwendraat. 2021. “Integrated Multi-Omics Analysis Reveals Common and Distinct Dysregulated Pathways for Genetic Subtypes of Frontotemporal Dementia.” bioRxiv. https://www.biorxiv.org/content/10.1101/2020.12.01.405894v2.abstract.

Mizutani, Akihiro, Yukiko Kuroda, Akira Futatsugi, Teiichi Furuichi, and Katsuhiko Mikoshiba. 2008. “Phosphorylation of Homer3 by Calcium/calmodulin-Dependent Kinase II Regulates a Coupling State of Its Target Molecules in Purkinje Cells.” The Journal of Neuroscience: The Official Journal of the Society for Neuroscience 28 (20): 5369–82.

Murley, Alexander G., Matthew A. Rouse, P. Simon Jones, Rong Ye, Frank H. Hezemans, Claire O’Callaghan, Polytimi Frangou, et al. 2020. “GABA and Glutamate Deficits from Frontotemporal Lobar Degeneration Are Associated with Disinhibition.” Brain: A Journal of Neurology 143 (11): 3449–62.

Nana, Alissa L., Manu Sidhu, Stephanie E. Gaus, Ji-Hye L. Hwang, Libo Li, Youngsoon Park, Eun-Joo Kim, et al. 2019. “Neurons Selectively Targeted in Frontotemporal Dementia Reveal Early Stage TDP-43 Pathobiology.” Acta Neuropathologica 137 (1): 27–46.

Neumann, M., D. M. Sampathu, L. K. Kwong, A. C. Truax, M. C. Micsenyi, T. T. Chou, J. Bruce, et al. 2006. “Ubiquitinated TDP-43 in Frontotemporal Lobar Degeneration and Amyotrophic Lateral Sclerosis.” Science 314 (5796): 130–33.

Patrick, Ellis, Mariko Taga, Ayla Ergun, Bernard Ng, William Casazza, Maria Cimpean, Christina Yung, et al. 2020. “Deconvolving the Contributions of Cell-Type Heterogeneity on Cortical Gene Expression.” PLoS Computational Biology 16 (8): e1008120.

Pidsley, Ruth, Chloe C. Y Wong, Manuela Volta, Katie Lunnon, Jonathan Mill, and Leonard C. Schalkwyk. 2013. “A Data-Driven Approach to Preprocessing Illumina 450K Methylation Array Data.” BMC Genomics 14 (May): 293.

Polymenidou, Magdalini, Clotilde Lagier-Tourenne, Kasey R. Hutt, Stephanie C. Huelga, Jacqueline Moran, Tiffany Y. Liang, Shuo-Chien Ling, et al. 2011. “Long Pre-mRNA Depletion and RNA Missplicing Contribute to Neuronal Vulnerability from Loss of TDP-43.” Nature Neuroscience. https://doi.org/10.1038/nn.2779.

Prudencio, Mercedes, Veronique V. Belzil, Ranjan Batra, Christian A. Ross, Tania F. Gendron, Luc J. Pregent, Melissa E. Murray, et al. 2015. “Distinct Brain Transcriptome Profiles in C9orf72-Associated and Sporadic ALS.” Nature Neuroscience 18 (8): 1175–82.

Ramesh, Geeta, Andrew G. MacLean, and Mario T. Philipp. 2013. “Cytokines and Chemokines at the Crossroads of Neuroinflammation, Neurodegeneration, and Neuropathic Pain.” Mediators of Inflammation 2013 (August): 480739.

Rempe, Ralf G., Anika M. S. Hartz, and Björn Bauer. 2016. “Matrix Metalloproteinases in the Brain and Blood-Brain Barrier: Versatile Breakers and Makers.” Journal of Cerebral Blood Flow and Metabolism: Official Journal of the International Society of Cerebral Blood Flow and Metabolism 36 (9): 1481–1507.

Renton, Alan E., Elisa Majounie, Adrian Waite, Javier Simón-Sánchez, Sara Rollinson, J. Raphael Gibbs, Jennifer C. Schymick, et al. 2011. “A Hexanucleotide Repeat Expansion in C9ORF72 Is the Cause of Chromosome 9p21-Linked ALS-FTD.” Neuron 72 (2): 257–68.

Rosa Ma, X., Mercedes Prudencio, Yuka Koike, Sarat C. Vatsavayai, Garam Kim, Fred Harbinski, Caitlin M. Rodriguez, et al. 2021. “TDP-43 Represses Cryptic Exon Inclusion in FTD/ALS Gene UNC13A.” bioRxiv. https://doi.org/10.1101/2021.04.02.438213.

Rot, Gregor, Zhen Wang, Ina Huppertz, Miha Modic, Tina Lenče, Martina Hallegger, Nejc Haberman, Tomaž Curk, Christian von Mering, and Jernej Ule. 2017. “High-Resolution RNA Maps Suggest Common Principles of Splicing and Polyadenylation Regulation by TDP-43.” Cell Reports 19 (5): 1056–67.

Ruegsegger, Céline, David M. Stucki, Silvio Steiner, Nico Angliker, Julika Radecke, Eva Keller, Benoît Zuber, Markus A. Rüegg, and Smita Saxena. 2016. “Impaired mTORC1-Dependent Expression of Homer-3 Influences SCA1 Pathophysiology.” Neuron 89 (1): 129–46.

Santillo, A. F., C. Nilsson, and E. Englund. 2013. “Von Economo Neurones Are Selectively Targeted in Frontotemporal Dementia.” Neuropathology and Applied Neurobiology 39 (5): 572–79.

Šarac, Helena, Marija žagar, Davorka VranjeŠ, Neven Henigsberg, Ervina Bilić, and Goran PavliŠa. 2008. “Magnetic Resonance Imaging and Magnetic Resonance Spectroscopy in a Patient with Amyotrophic Lateral Sclerosis and Frontotemporal Dementia.” Collegium Antropologicum 32 (1): 205–10.

Schroeder, Andreas, Odilo Mueller, Susanne Stocker, Ruediger Salowsky, Michael Leiber, Marcus Gassmann, Samar Lightfoot, Wolfram Menzel, Martin Granzow, and Thomas Ragg. 2006. “The RIN: An RNA Integrity Number for Assigning Integrity Values to RNA Measurements.” BMC Molecular Biology 7 (January): 3.

Skene, Nathan G., and Seth G. N. Grant. 2016. “Identification of Vulnerable Cell Types in Major Brain Disorders Using Single Cell Transcriptomes and Expression Weighted Cell Type Enrichment.” Frontiers in Neuroscience 10 (January): 16.

Subramanian, Aravind, Pablo Tamayo, Vamsi K. Mootha, Sayan Mukherjee, Benjamin L. Ebert, Michael A. Gillette, Amanda Paulovich, et al. 2005. “Gene Set Enrichment Analysis: A Knowledge-Based Approach for Interpreting Genome-Wide Expression Profiles.” Proceedings of the National Academy of Sciences of the United States of America 102 (43): 15545–50.

Tam, Oliver H., Nikolay V. Rozhkov, Regina Shaw, Duyang Kim, Isabel Hubbard, Samantha Fennessey, Nadia Propp, et al. 2019. “Postmortem Cortex Samples Identify Distinct Molecular Subtypes of ALS: Retrotransposon Activation, Oxidative Stress, and Activated Glia.” Cell Reports 29 (5): 1164–77.e5.

Tan, Rachel H., Emma Devenney, Carol Dobson-Stone, John B. Kwok, John R. Hodges, Matthew C. Kiernan, Glenda M. Halliday, and Michael Hornberger. 2014. “Cerebellar Integrity in the Amyotrophic Lateral Sclerosis-Frontotemporal Dementia Continuum.” PloS One 9 (8): e105632.

Tian, Yuan, Tiffany J. Morris, Amy P. Webster, Zhen Yang, Stephan Beck, Andrew Feber, and Andrew E. Teschendorff. 2017. “ChAMP: Updated Methylation Analysis Pipeline for Illumina BeadChips.” Bioinformatics 33 (24): 3982–84.

Tollervey, James R., Tomaž Curk, Boris Rogelj, Michael Briese, Matteo Cereda, Melis Kayikci, Julian Konig, et al. 2011. “Characterizing the RNA Targets and Position-Dependent Splicing Regulation by TDP-43.” Nature Neuroscience 14 (4): 452–58.

Vogt, Miriam A., Zahra Ehsaei, Philip Knuckles, Adrian Higginbottom, Michaela S. Helmbrecht, Tilo Kunath, Kevin Eggan, et al. 2018. “TDP-43 Induces p53-Mediated Cell Death of Cortical Progenitors and Immature Neurons.” Scientific Reports 8 (1): 8097.

Wang, Xuran, Jihwan Park, Katalin Susztak, Nancy R. Zhang, and Mingyao Li. 2019. “Bulk Tissue Cell Type Deconvolution with Multi-Subject Single-Cell Expression Reference.” Nature Communications 10 (1): 380.

Woollacott, Ione O. C., Christina E. Toomey, Catherine Strand, Robert Courtney, Bridget C. Benson, Jonathan D. Rohrer, and Tammaryn Lashley. 2020. “Microglial Burden, Activation and Dystrophy Patterns in Frontotemporal Lobar Degeneration.” Journal of Neuroinflammation 17 (1): 234.

Yousef, Ahmed, John L. Robinson, David J. Irwin, Matthew D. Byrne, Linda K. Kwong, Edward B. Lee, Yan Xu, et al. 2017. “Neuron Loss and Degeneration in the Progression of TDP-43 in Frontotemporal Lobar Degeneration.” Acta Neuropathologica Communications 5 (1): 68.

Yu, Guangchuang, Li-Gen Wang, Yanyan Han, and Qing-Yu He. 2012. “clusterProfiler: An R Package for Comparing Biological Themes among Gene Clusters.” Omics: A Journal of Integrative Biology 16 (5): 284–87.

Zamanian, Jennifer L., Lijun Xu, Lynette C. Foo, Navid Nouri, Lu Zhou, Rona G. Giffard, and Ben A. Barres. 2012. “Genomic Analysis of Reactive Astrogliosis.” The Journal of Neuroscience: The Official Journal of the Society for Neuroscience 32 (18): 6391–6410.

Zhang, Jiasheng, Dmitry Velmeshev, Kei Hashimoto, Yu-Hsin Huang, Jeffrey W. Hofmann, Xiaoyu Shi, Jiapei Chen, et al. 2020. “Neurotoxic Microglia Promote TDP-43 Proteinopathy in Progranulin Deficiency.” Nature 588 (7838): 459–65.

