## Supplementary Figures 1-18 for "Transcriptomic analysis of frontotemporal lobar degeneration with TDP-43 pathology reveals cellular alterations across multiple brain regions"

Supplementary Figure 1: Covariate modelling. **3**

Supplementary Figure 2: Correlation matrices for technical factors. **4**

Supplementary Figure 3: Modelling covariates for differential expression. **5**

Supplementary Figure 4: Case-Control differential expression compared to correlations with PMI. **6**

Supplementary Figure 5: Comparing effect of C9orf72 mutation carriers on FTLD-TDP vs control differential expression. **7**

Supplementary Figure 6: Full pathway enrichment results. **8**

Supplementary Figure 7: Expression-fold changes of matrix metalloproteinase genes. **9**

Supplementary Figure 8: Expression-weighted cell-type enrichment (EWCE) analysis in the three brain regions. **10**

Supplementary Figure 9: Expression-fold changes of von Economo neuron marker genes. **11**

Supplementary Figure 10 : Partial overlaps between glia and glial activation gene lists. **13**

Supplementary Figure 11: Deconvolution results using an alternate cell-type reference and two different algorithms. **14**

Supplementary Figure 12: Comparing deconvolution estimates between dtangle and MuSiC for each cell type. **15**

Supplementary Figure 13: Comparing cell type proportion estimates between the Darmanis and Mathys reference panels for the shared cell-types. **16**

Supplementary Figure 14: Full results for correlations between microscopic atrophy scores and estimated cell-type proportions (from the Mathys reference panel). **17**

Supplementary Figure 15: Full results for correlations between macroscopic atrophy scores and estimated cell-type proportions (from the Mathys reference panel). **18**

Supplementary Figure 16: Correlations between gene expression and DNA methylation estimates of neuronal proportion. **19**

Supplementary Figure 17: Full results for correlations between microglia counts and estimated microglia proportions (from the Mathys reference panel). **20**

Supplementary Figure 18: Comparisons with TDP-43 knockdown genes from Brown et al. **21**

| | Covariate | Number of genes with improved BIC |
|---|---|---|
| **Frontal Cortex** | median 3' bias | 3120 |
| | % chimeras | 2001 |
| | % R1 transcript strand reads | 1675 |
| | % R2 transcript strand reads | 1647 |
| | mean read length | 819 |
| | % UTR bases | 779 |
| | % intergenic bases | 633 |
| | median 5' bias | 588 |
| | % coding bases | 558 |
| | % ribosomal bases | 444 |
| **Temporal Cortex** | % chimeras | 4584 |
| | % R2 transcript strand reads | 1415 |
| | median 3' bias | 1291 |
| | % R1 transcript strand reads | 1090 |
| | % pf reads aligned | 873 |
| | % intergenic bases | 821 |
| | % intronic bases | 667 |
| | mean read length | 635 |
| | % UTR bases | 616 |
| | % ribosomal bases | 489 |
| **Cerebellum** | median 3' bias | 2152 |
| | % coding bases | 1733 |
| | mean read length | 1286 |
| | % R2 transcript strand reads | 1236 |
| | median 5' bias | 1184 |
| | % R1 transcript strand reads | 1149 |
| | % UTR bases | 892 |
| | % chimeras | 633 |
| | % intergenic bases | 487 |
| | % ribosomal bases | 483 |

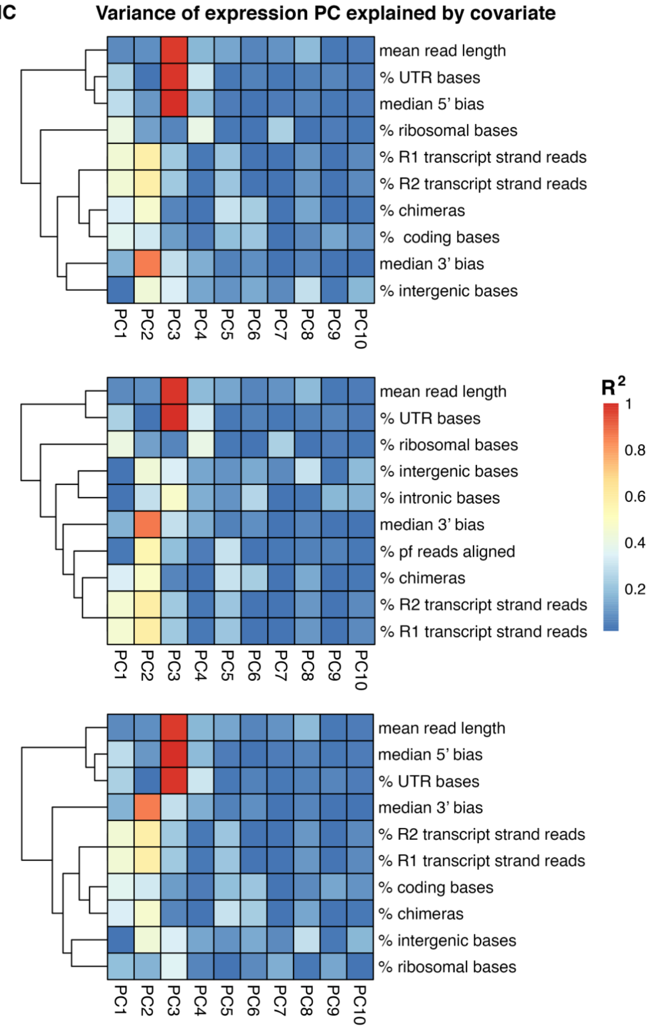


**Supplementary Figure 1: Covariate modelling.**
Left panels: Technical factors generated by PicardTools on the BAM files of all control and FTLD-TDP samples. For each tissue, each technical covariate was added to a base model (~disease) for differential gene expression. The number of genes with improved Bayesian Information Criteria (BIC) compared to the base model for the top 10 technical covariates is listed for each tissue. Right Panels: Linear regression model fits for the top 10 principal components of gene expression with a set of technical covariates in each tissue .Goodness of fit is represented by the $R^2$ value.

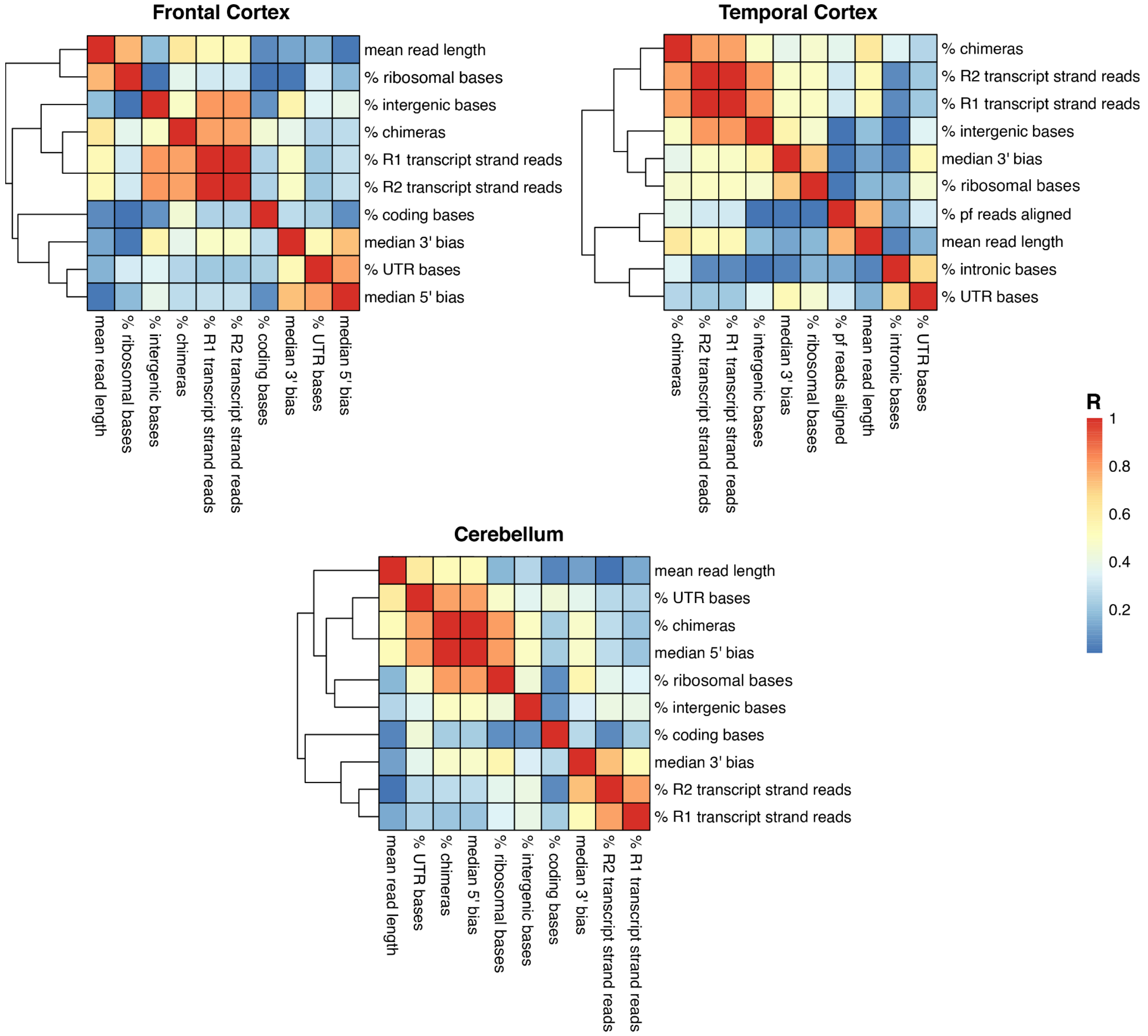


**Supplementary Figure 2: Correlation matrices for technical factors.**
Correlation matrices of the top 10 technical covariates for each tissue. Colored tiles correspond to Pearson correlation coefficients. Covariates for differential expression modeling were chosen such that highly correlated variables (R > 0.5) were excluded.

**a**

| | Model | Covariates | Number of genes with improved BIC |
|---|---|---|---|
| Frontal Cortex | 1 | ~ disease + sex + age | 2403 |
| | 2 | ~ disease + sex + age + median 3' bias | 3906 |
| | 3 | ~ disease + sex + age + median 3' bias + % coding bases | 2732 |
| | 4 | ~ disease + sex + age + median 3' bias + % coding bases + % ribosomal bases | 1932 |
| | 5 | ~ disease + sex + age + % R1 transcript strand reads | 2601 |
| | 6 | ~ disease + sex + age + % R1 transcript strand reads + % UTR bases | 1092 |
| | 7 | ~ disease + sex + age + % R1 transcript strand reads + % UTR bases + % coding bases | 408 |
| | 8 | ~ disease + sex + age + % R1 transcript strand reads + % UTR bases + % coding bases + total counts | 323 |
| Temporal Cortex | 1 | ~ disease + sex + age | 3242 |
| | 2 | ~ disease + sex + age + median 3' bias | 1594 |
| | 3 | ~ disease + sex + age + median 3' bias + % intronic bases | 1737 |
| | 4 | ~ disease + sex + age + median 3' bias + % intronic bases + % ribosomal bases | 46 |
| | 5 | ~ disease + sex + age + % R2 transcript strand reads | 1845 |
| | 6 | ~ disease + sex + age + % R2 transcript strand reads + % intronic bases | 2374 |
| | 7 | ~ disease + sex + age + % R2 transcript strand reads + % intronic bases + % ribosomal bases | 4225 |
| Cerebellum | 1 | ~ disease + sex + age | 3312 |
| | 2 | ~ disease + sex + age + median 3' bias | 2449 |
| | 3 | ~ disease + sex + age + median 3' bias + coding bases | 3030 |
| | 4 | ~ disease + sex + age + % R2 transcript strand reads | 3695 |
| | 5 | ~ disease + sex + age + % R2 transcript strand reads + median 5' bias | 2128 |
| | 6 | ~ disease + sex + age + % R2 transcript strand reads + median 5' bias + % ribosomal bases | 452 |

**b**

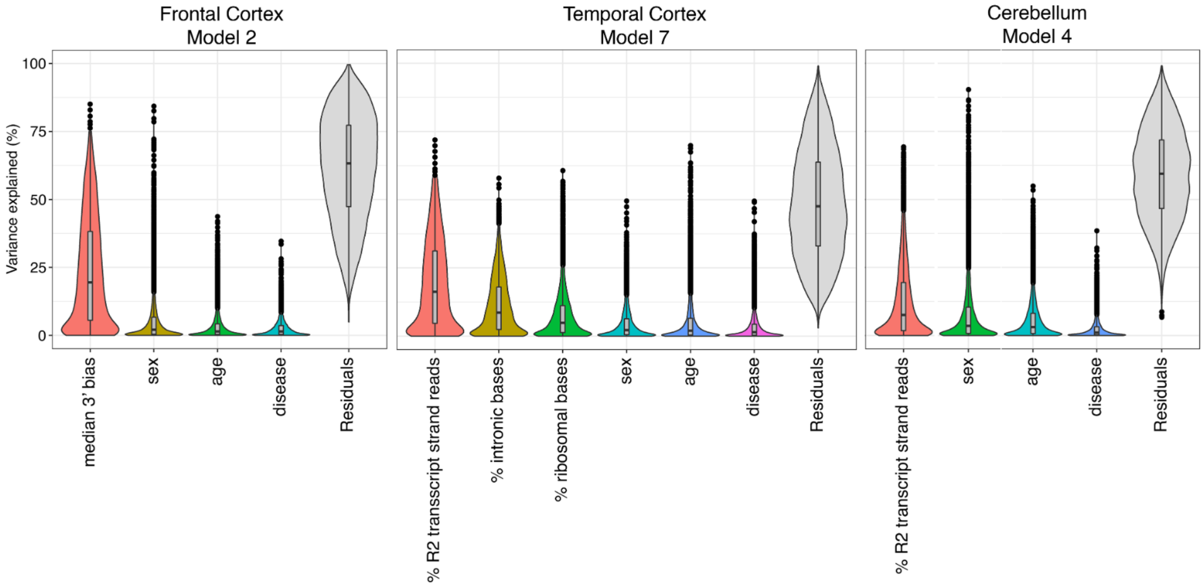


**Supplementary Figure 3: Modelling covariates for differential expression.**
a) Each differential expression model fitted, with number of genes with improved Bayesian Information Criterion (BIC) compared to the base model (~ disease). Model 2 (frontal cortex), model 7 (temporal cortex), and model 4 (cerebellum) were chosen for differential gene expression analysis as they provided the best BIC improvement . b) VariancePartition plots for the models with the best BIC improvement. On average, technical factors explain a greater proportion of variance than clinical factors.

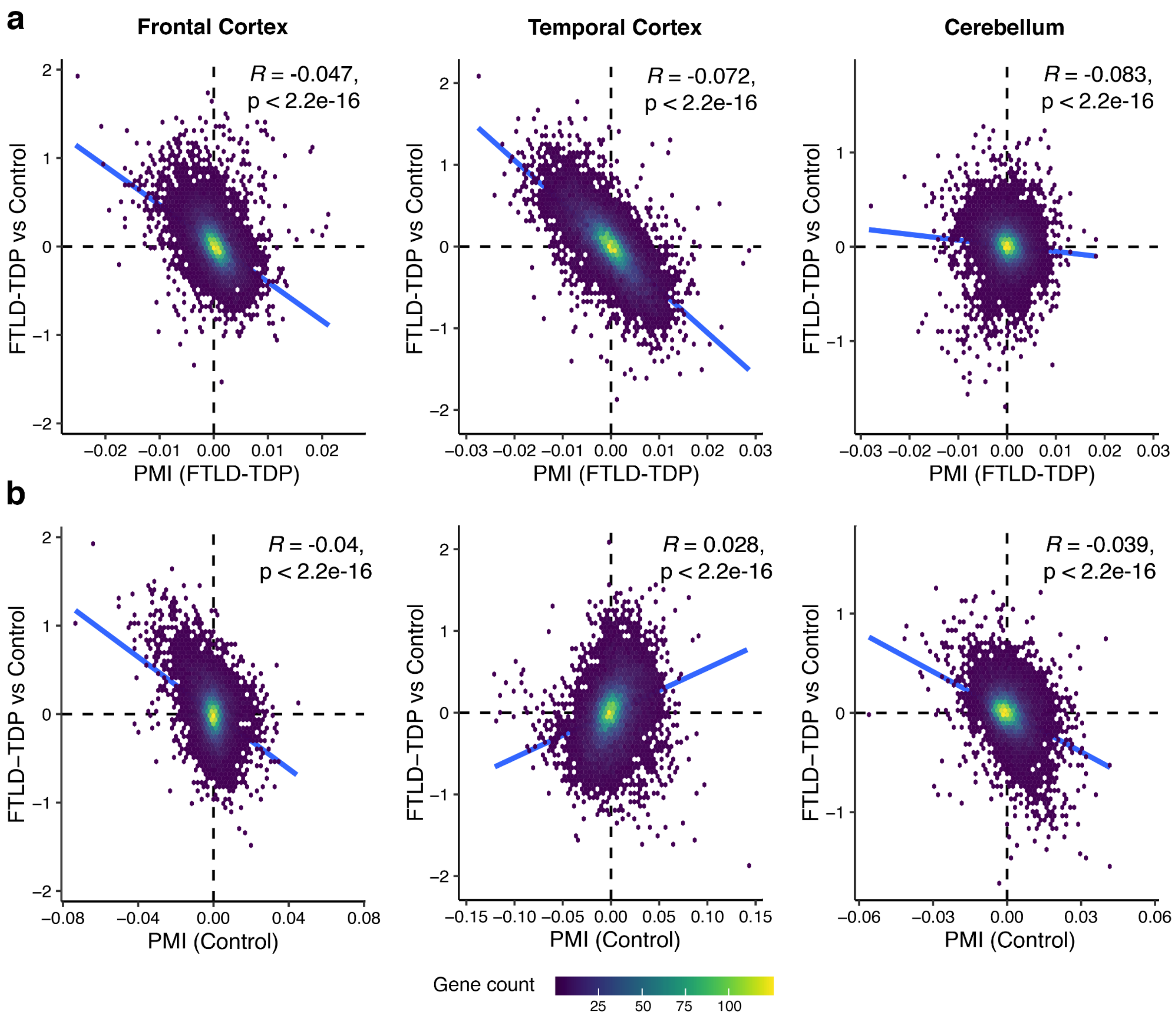


**Supplementary Figure 4: Case-Control differential expression compared to correlations with PMI.**

Scatter plots showing the relationships between FTLD-TDP vs Control differential expression effect size ($log_2$ fold change) for each gene, compared with effect sizes of correlation with PMI in FTLD-TDP samples only (a) or control samples only (b). Individual points are genes, color refers to density of overlapping points. R refers to the Pearson correlation coefficient.

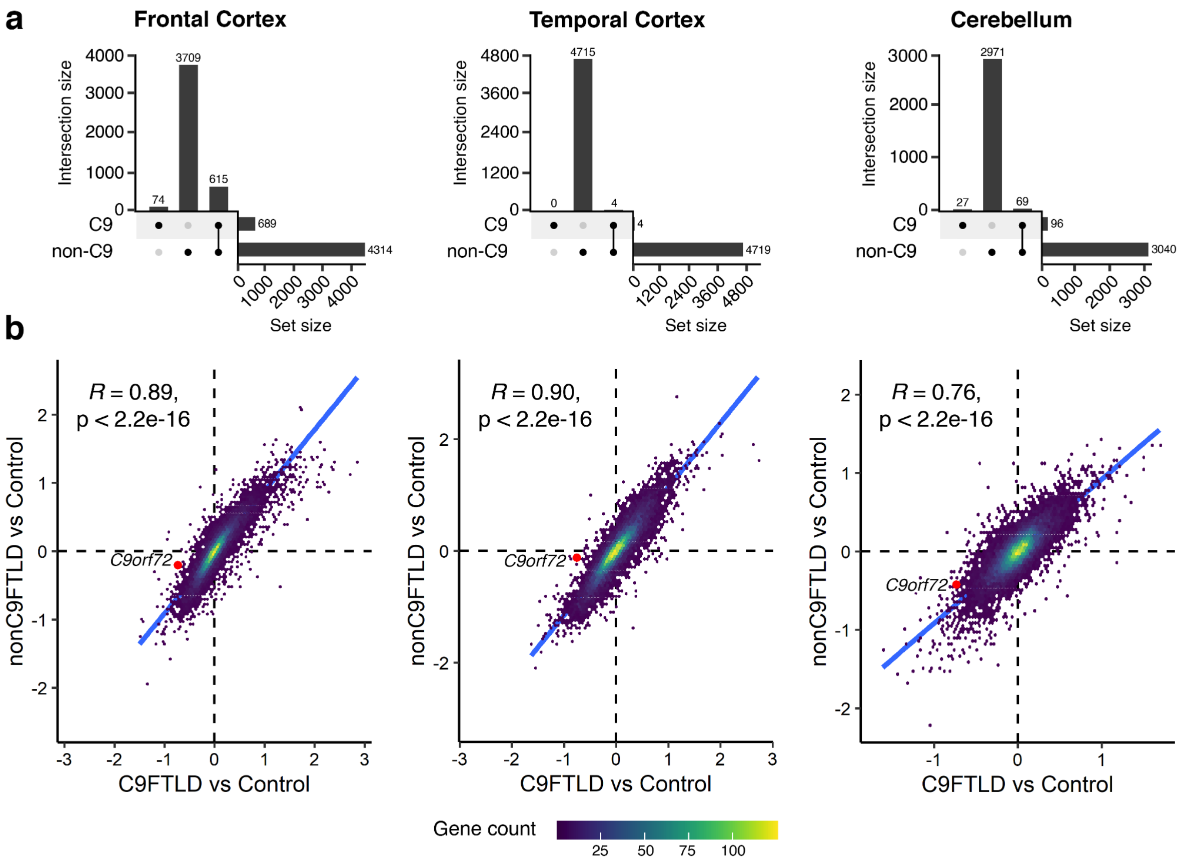


**Supplementary Figure 5: Comparing effect of C9orf72 mutation carriers on FTLD-TDP vs control differential expression.**

a) Upset plots showing the number of distinct and overlapping differentially expressed genes from the C9orf72 vs Control and non-C9orf72 vs Control comparisons. b) Scatter plots show correlation between $\log_2$ fold changes per change between the C9orf72-FTLD-TDP samples and controls (x axis) and the non-C9orf72 FTLD-TDP samples and controls (y axis). *C9orf72* gene highlighted in red. Individual points are genes, color refers to density of overlapping points. R refers to the Pearson correlation coefficient.

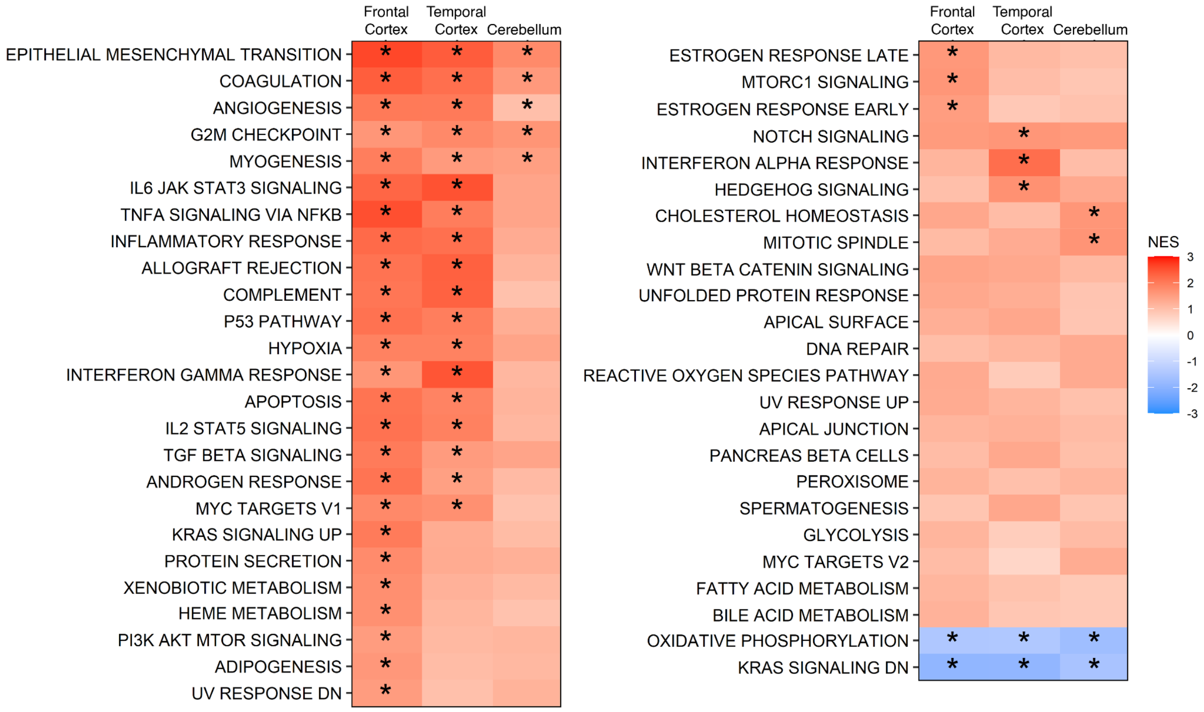


**Supplementary Figure 6: Full pathway enrichment results.**

GSEA results for all molecular signatures database (mSigDB) pathways. Colored tiles represent the normalized-enrichment score (NES) for each pathway. Asterisks refer to Bonferroni-adjusted P-values less than 0.05.

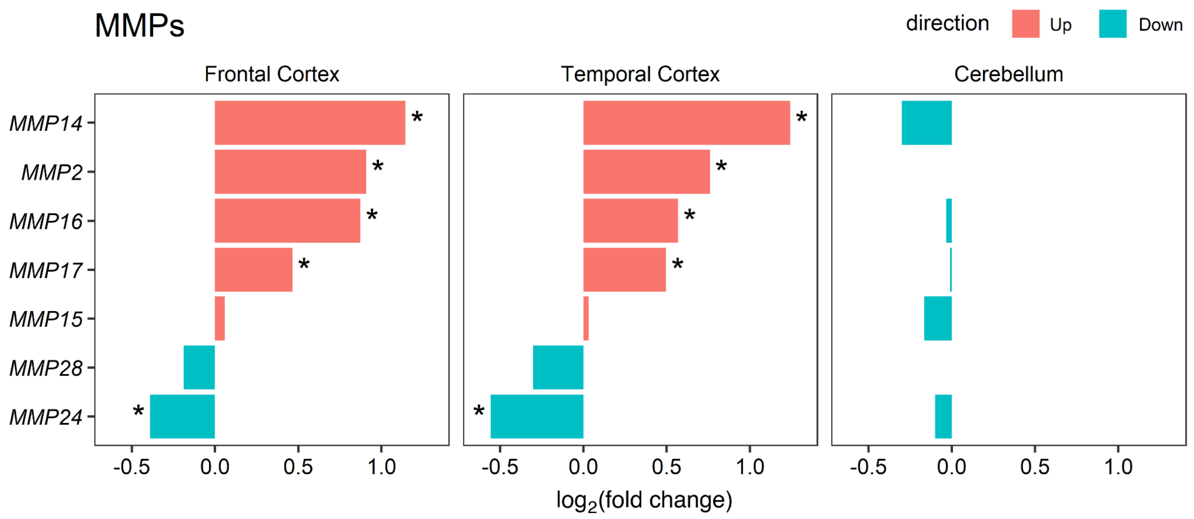


**Supplementary Figure 7: Expression-fold changes of matrix metalloproteinase genes.**

Barplots show the $\log_2$-fold changes of matrix metalloproteinase (MMP) genes in each region from differential expression. Asterisks refer to Bonferroni-adjusted P-values less than 0.05. Most MMP genes were differentially upregulated in the cortices.

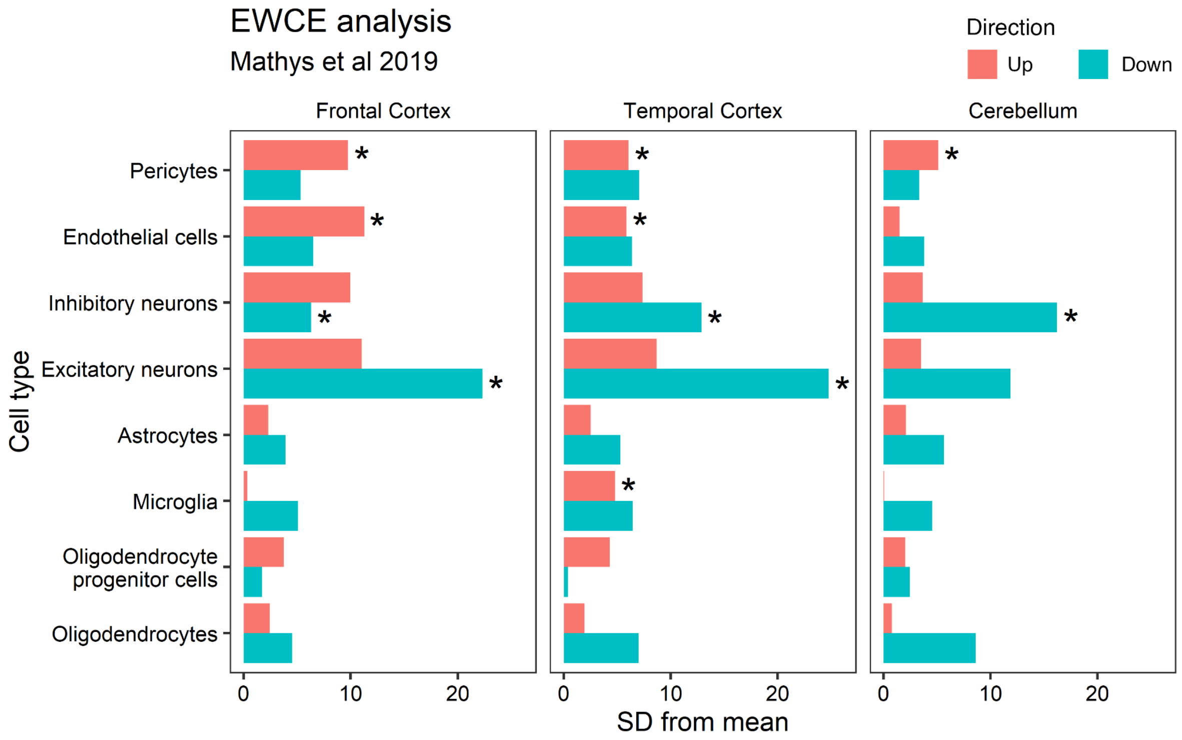


**Supplementary Figure 8: Expression-weighted cell-type enrichment (EWCE) analysis in the three brain regions.**
Cell-type specificity data generated from human frontal cortex data (Mathys et al, 2019). Enrichment of upregulated genes (red) and downregulated genes (blue) in cell-type specific gene sets shown as number of standard deviations from the mean of 1000 permutations. Asterisks refers to adjusted P-values less than 0.05.

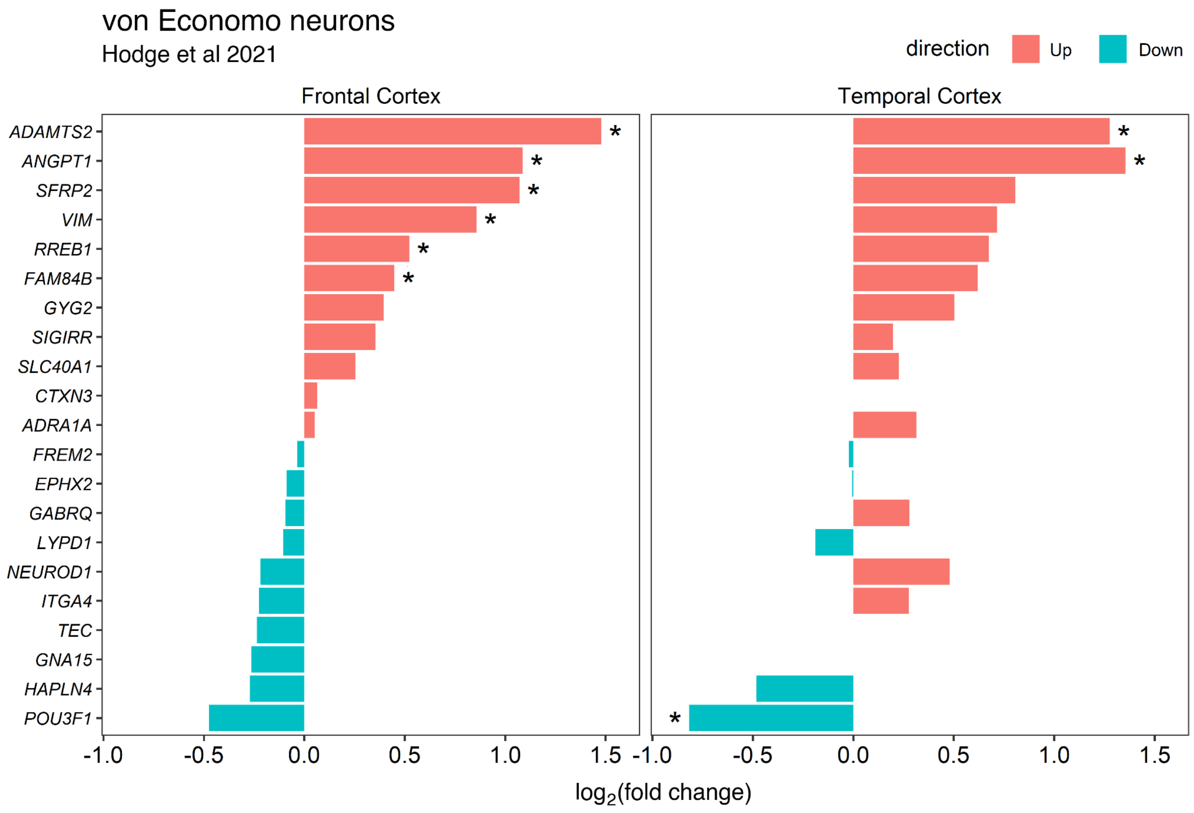


**Supplementary Figure 9: Expression-fold changes of von Economo neuron marker genes.**

Marker genes for von Economo neurons (VENs) were defined via single-cell RNA-seq profiling of the human frontoinsular cortex (Hodge et al, 2021). Bar plots show the $\log_2$-fold changes of the marker genes from differential expression in the frontal and temporal regions. Asterisks refer to Bonferroni-adjusted P-values less than 0.05. Contrary to the selective vulnerability of VENs, VEN-specific genes were mainly upregulated, with the upregulated genes (red) exhibiting larger effect sizes than the downregulated genes (blue).

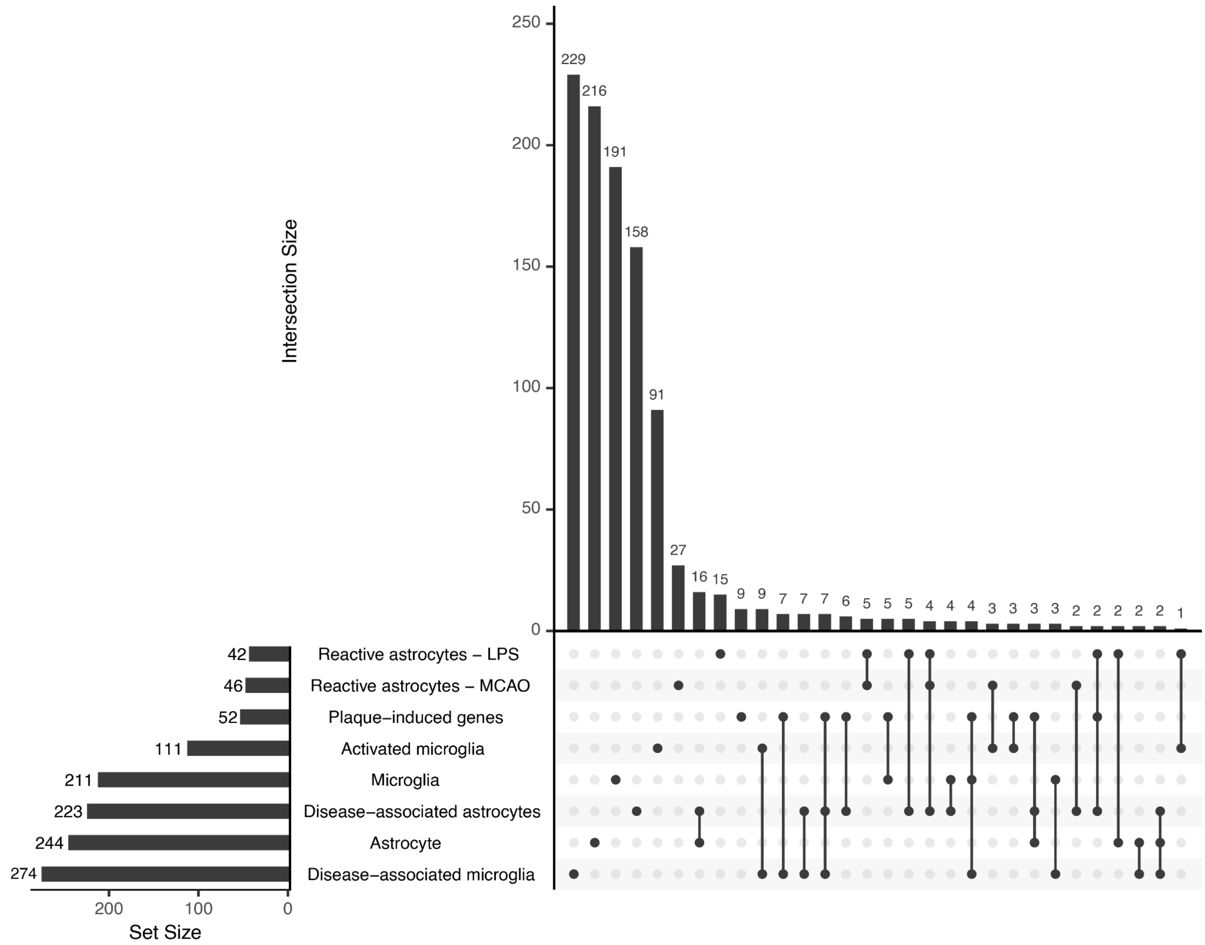


**Supplementary Figure 10 : Partial overlaps between glia and glial activation gene lists.** Upset plot comparing all glia and glial activation gene lists used in enrichment analysis. Shown are the total number of genes for each set (horizontal bars) and the number of exclusive or overlapping genes (vertical bars). The sets are largely distinct, with the exception of plaque-induced genes (only 9 of the 52 genes are unique to this set).

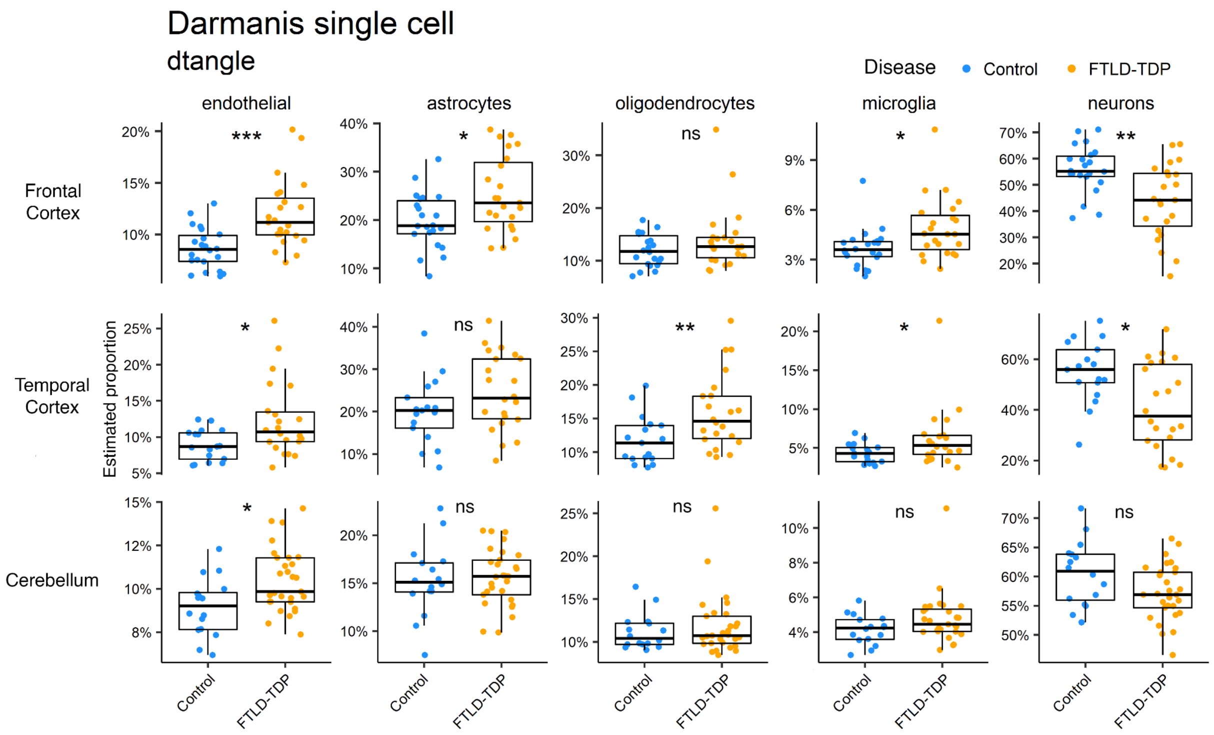


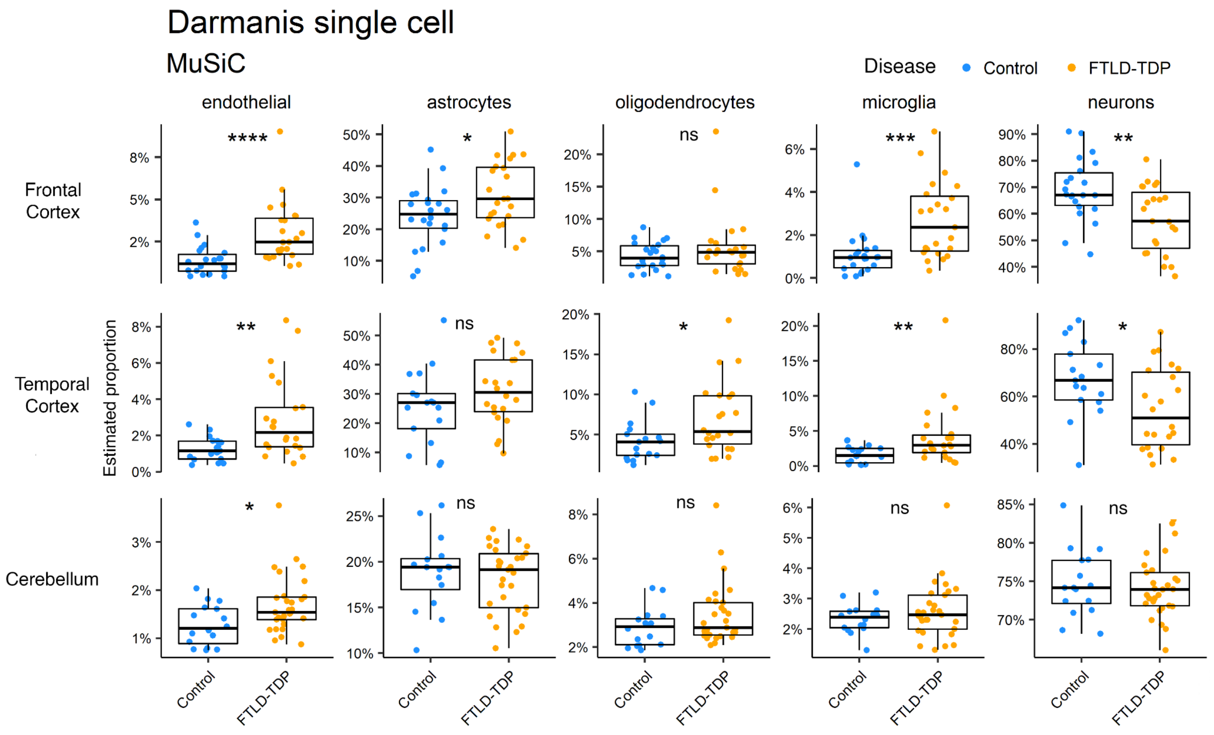


**Supplementary Figure 11: Deconvolution results using an alternate cell-type reference and two different algorithms.**

Top panel: Purified single cell RNA-seq profiles from Darmanis et al, 2015, applied with the dtangle algorithm. Bottom panel: Darmanis profiles applied to the MuSIC algorithm. Asterisks in both panels represent Bonferroni-adjusted P-values from Wilcoxon rank sum tests comparing cell-type estimates between FTLD-TDP and control samples. *** $p < 1e-4$; ** $p < 1e-3$; * $p < 0.05$; ns $p > 0.05$.

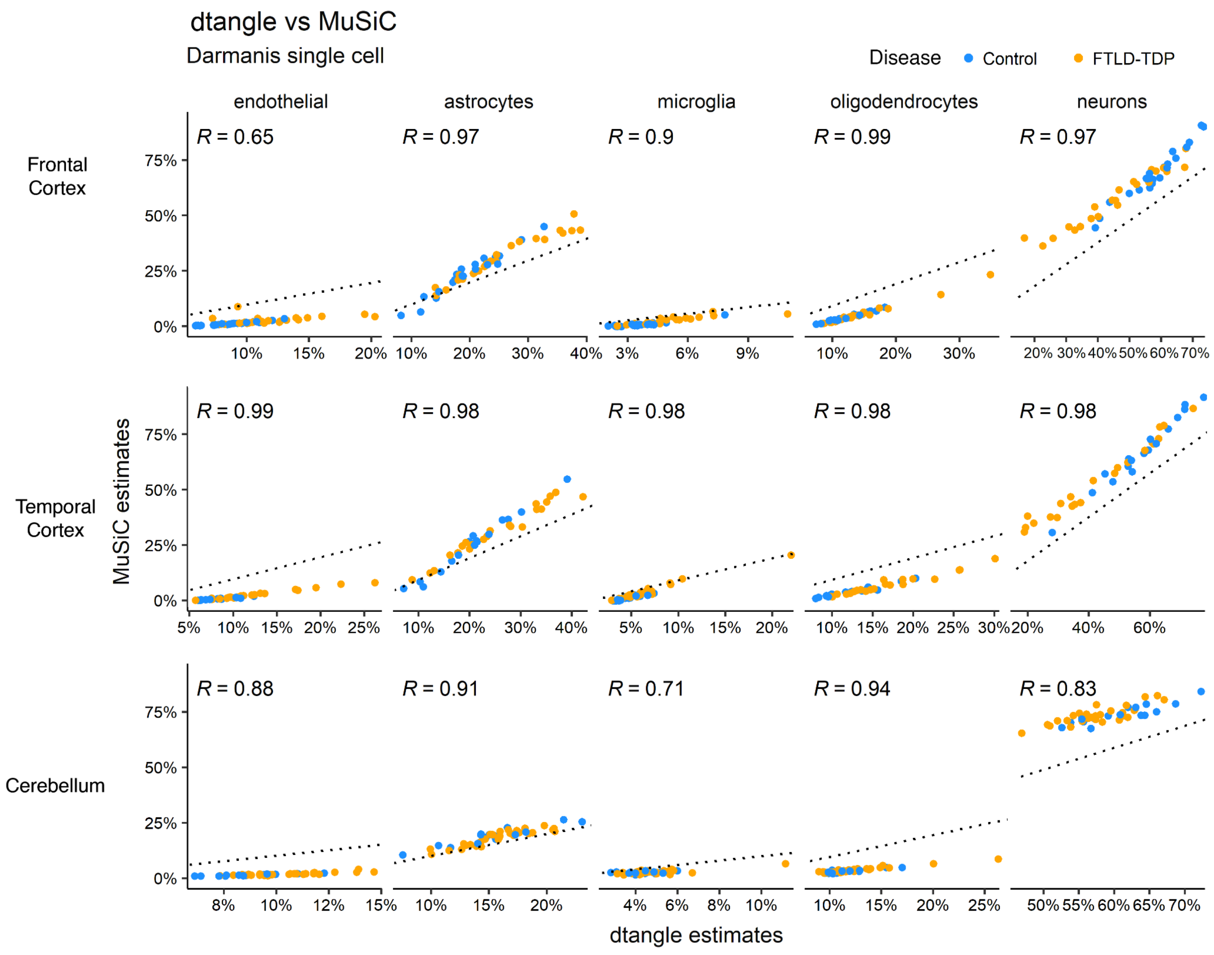


**Supplementary Figure 12: Comparing deconvolution estimates between dtangle and MuSiC for each cell type.**

Comparing decovonolution estimates between the  MuSiC and dtangle algorithms using the Darmanis et al single cell RNA-seq reference data. Dotted line is x=y. R refers to the Spearman correlation coefficient. The estimated proportions from MuSiC and dtangle are highly correlated, although the magnitude of the estimates differ considerably.

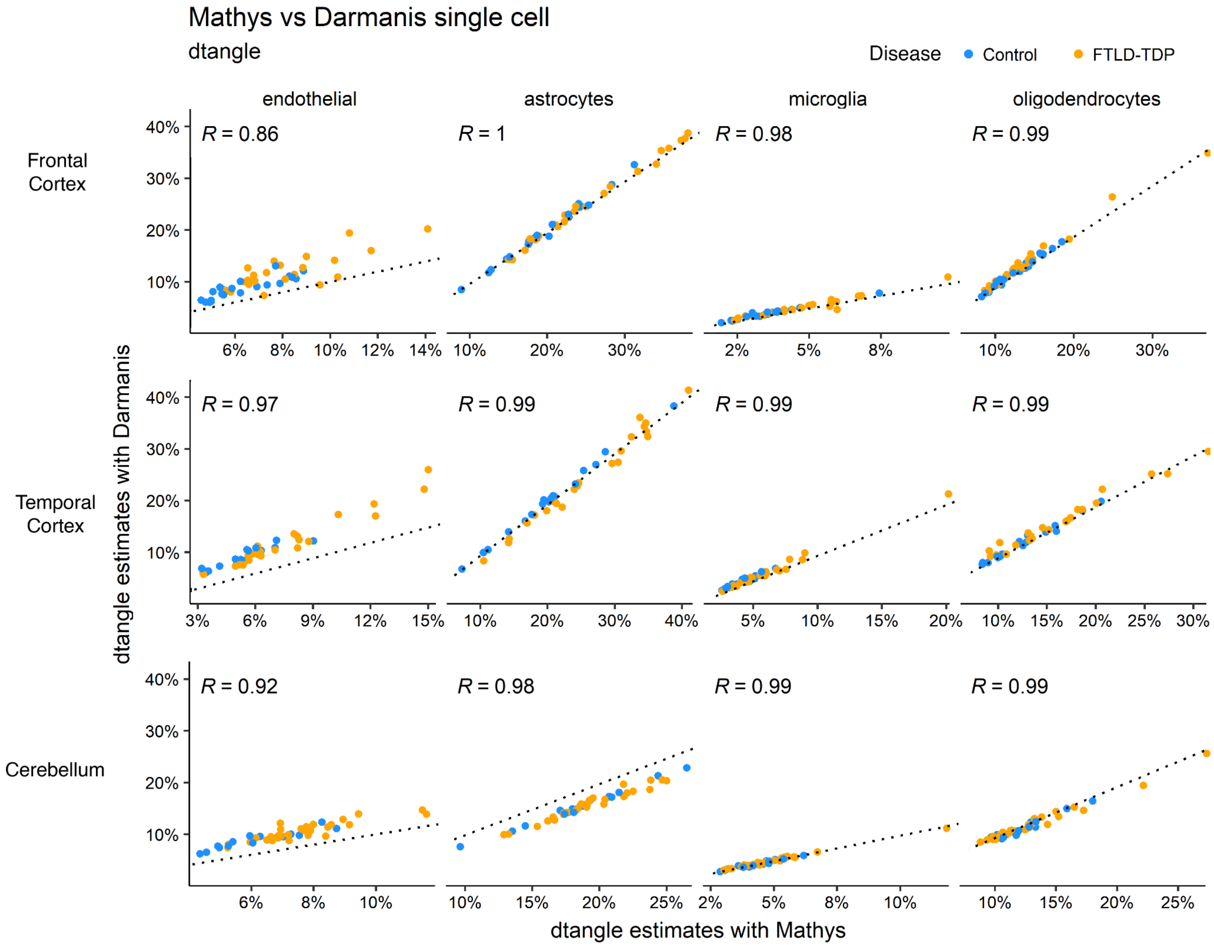


**Supplementary Figure 13: Comparing cell type proportion estimates between the Darmanis and Mathys reference panels for the shared cell-types.**

Comparing deconvolution estimates from dtangle between the Darmanis and Mathys reference panels. Dotted line is x=y. R refers to the Spearman correlation coefficient. Estimates from both reference panels are strongly correlated in the four overlapping cell-types.

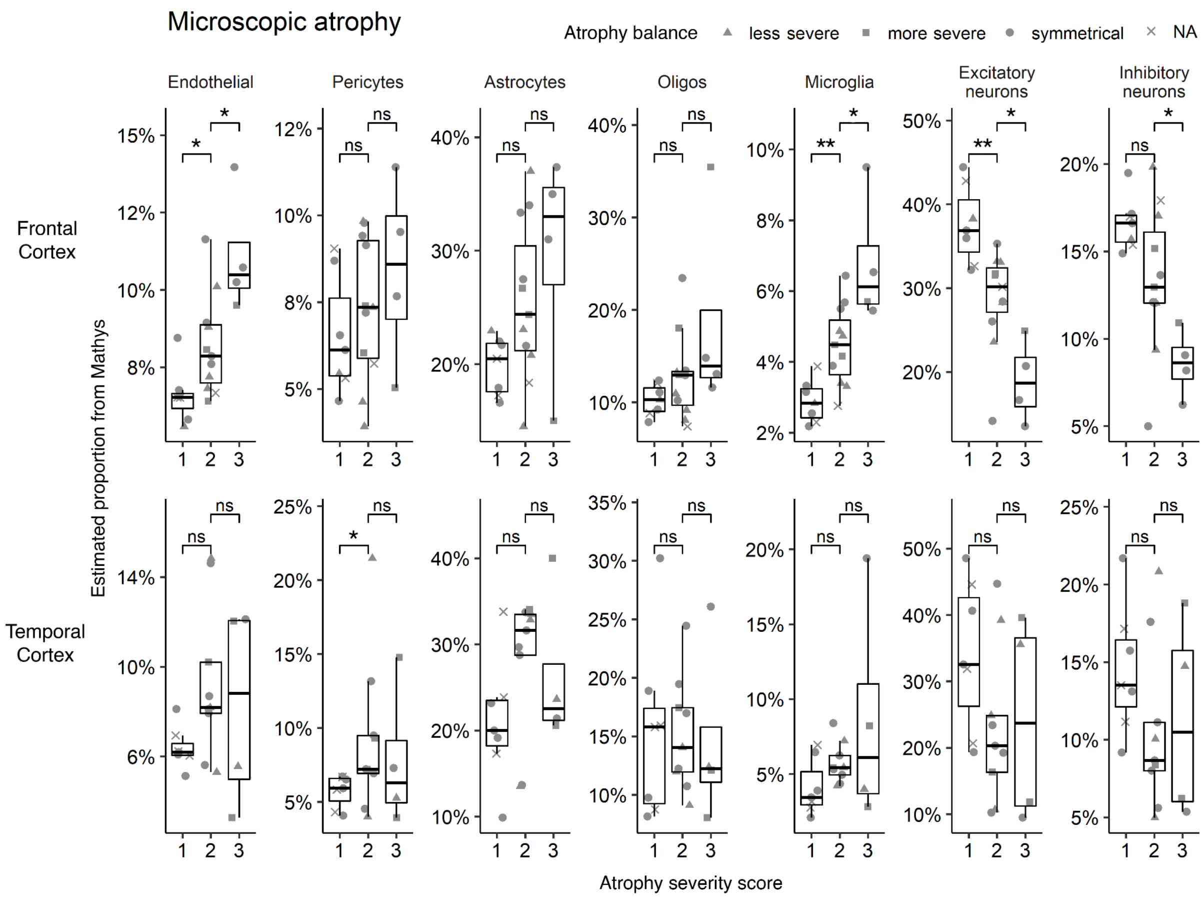


**Supplementary Figure 14: Full results for correlations between microscopic atrophy scores and estimated cell-type proportions (from the Mathys reference panel).** Samples highlighted by whether microscopic atrophy was judged to be symmetric between both hemispheres, less, or more severe on the side used for sequencing. Cell-type proportion estimates for the FTLD-TDP samples were compared between all atrophy stages using the Wilcoxon rank sum test. P-values were Bonferroni-corrected for multiple-testing. *** p < 1e-4; ** p < 1e-3; * p < 0.05; ns p > 0.05.

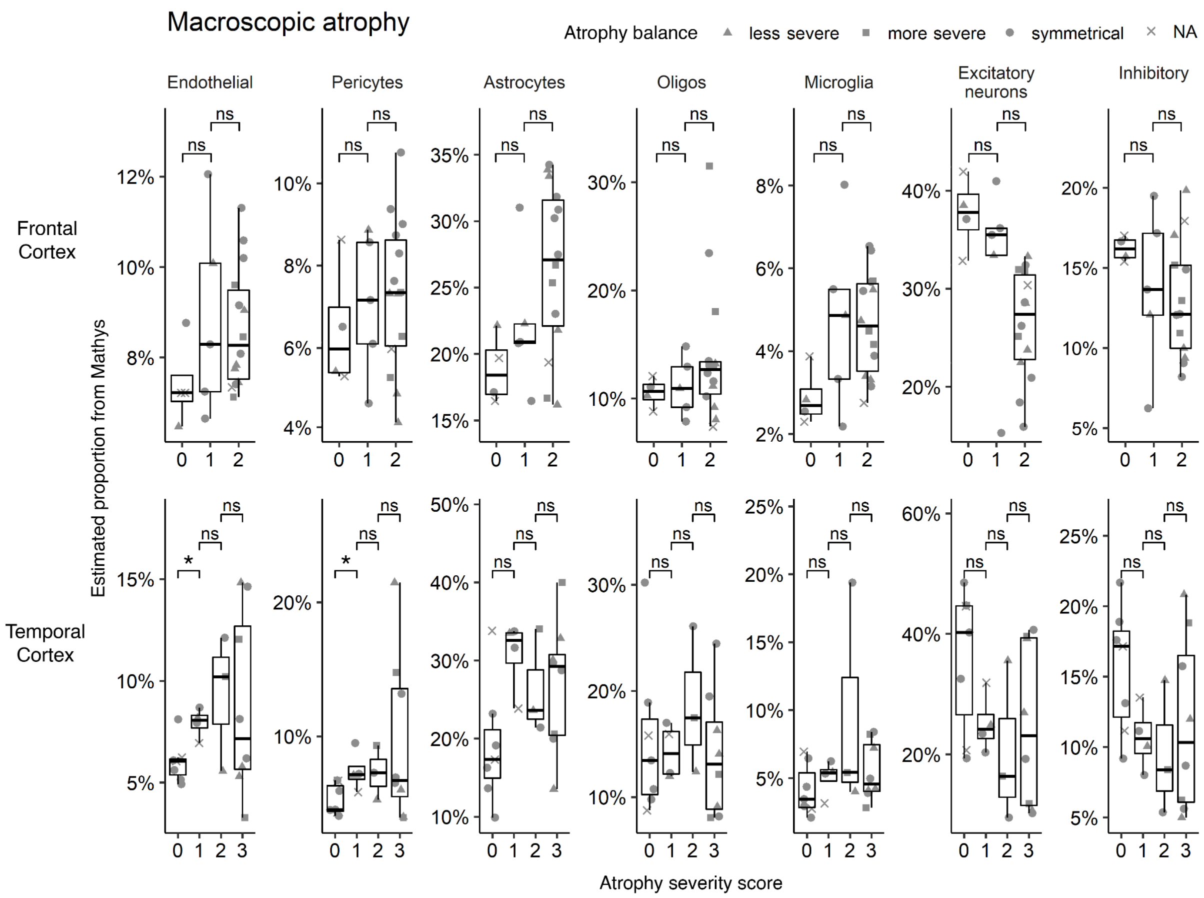


**Supplementary Figure 15: Full results for correlations between macroscopic atrophy scores and estimated cell-type proportions (from the Mathys reference panel).** Samples highlighted by whether macroscopic atrophy was judged to be symmetric between both hemispheres, less, or more severe on one side. Cell-type proportion estimates for the FTLD-TDP samples were compared between all atrophy stages using the Wilcoxon rank sum test. P-values were Bonferroni-corrected for multiple-testing. *** $p < 1e-4$; ** $p < 1e-3$; * $p < 0.05$; ns $p > 0.05$.

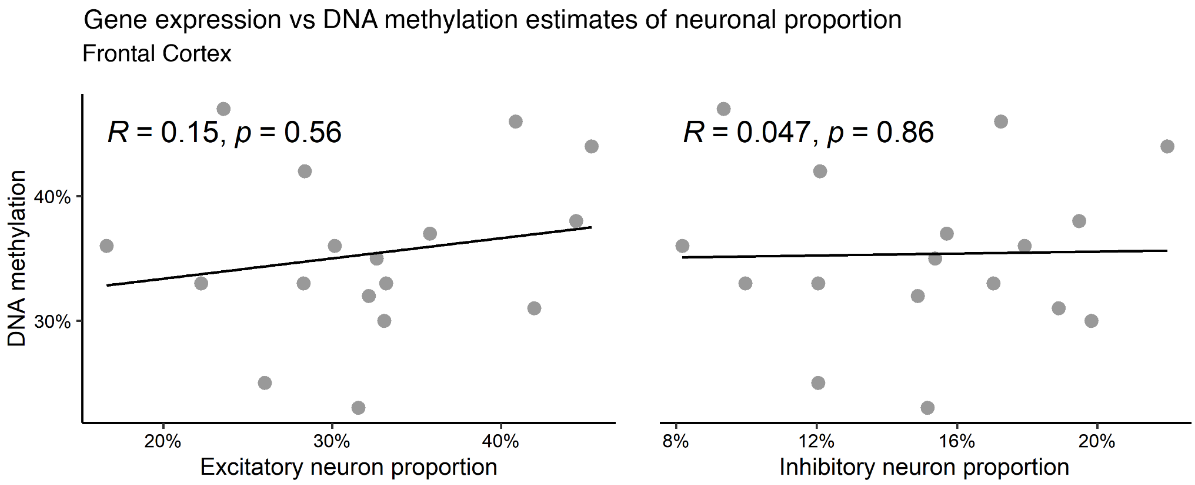


**Supplementary Figure 16: Correlations between gene expression and DNA methylation estimates of neuronal proportion.**

Scatter plots show Spearman correlations between neuronal proportions from DNA methylation and excitatory and inhibitory neuron estimates derived from gene expression using the Mathys reference panel. Only matching frontal cortex samples were correlated. No significant association ($P < 0.05$) was observed between the two measures.

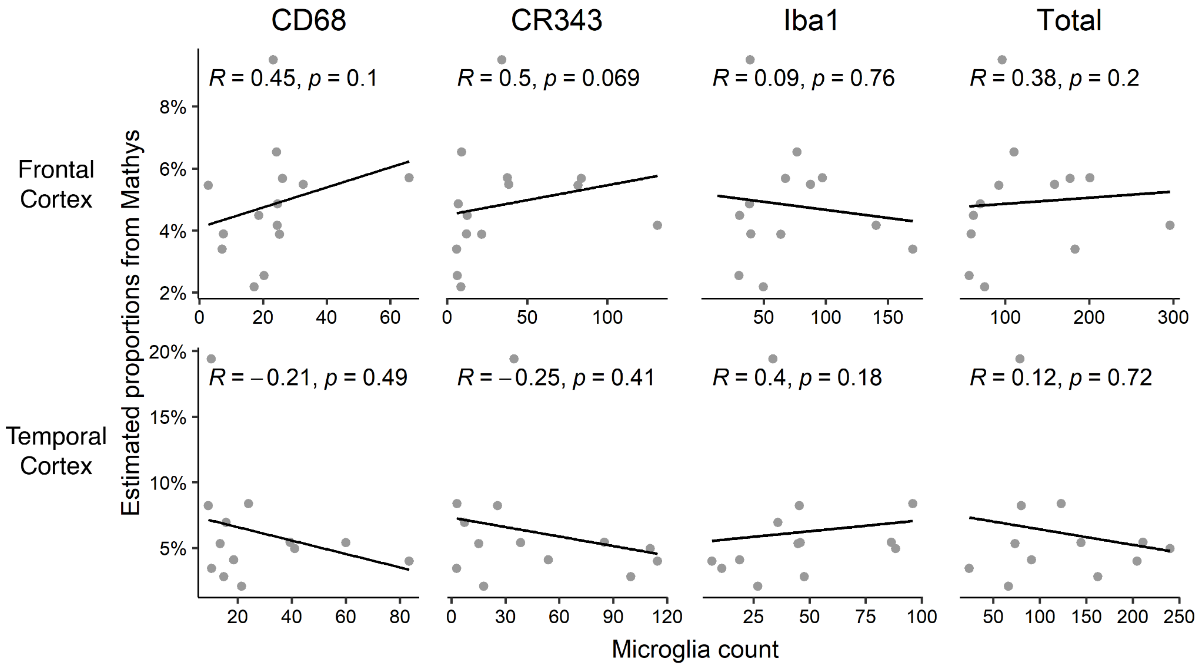


**Supplementary Figure 17: Full results for correlations between microglia counts and estimated microglia proportions (from the Mathys reference panel).**

Scatter plots show Spearman correlations between microglia counts of each marker (CD68, CR343, and IBA1) and estimated microglia proportions from the Mathys reference panel. No measure significantly correlated ($P < 0.05$) with microglia proportion in either the frontal or temporal cortex.

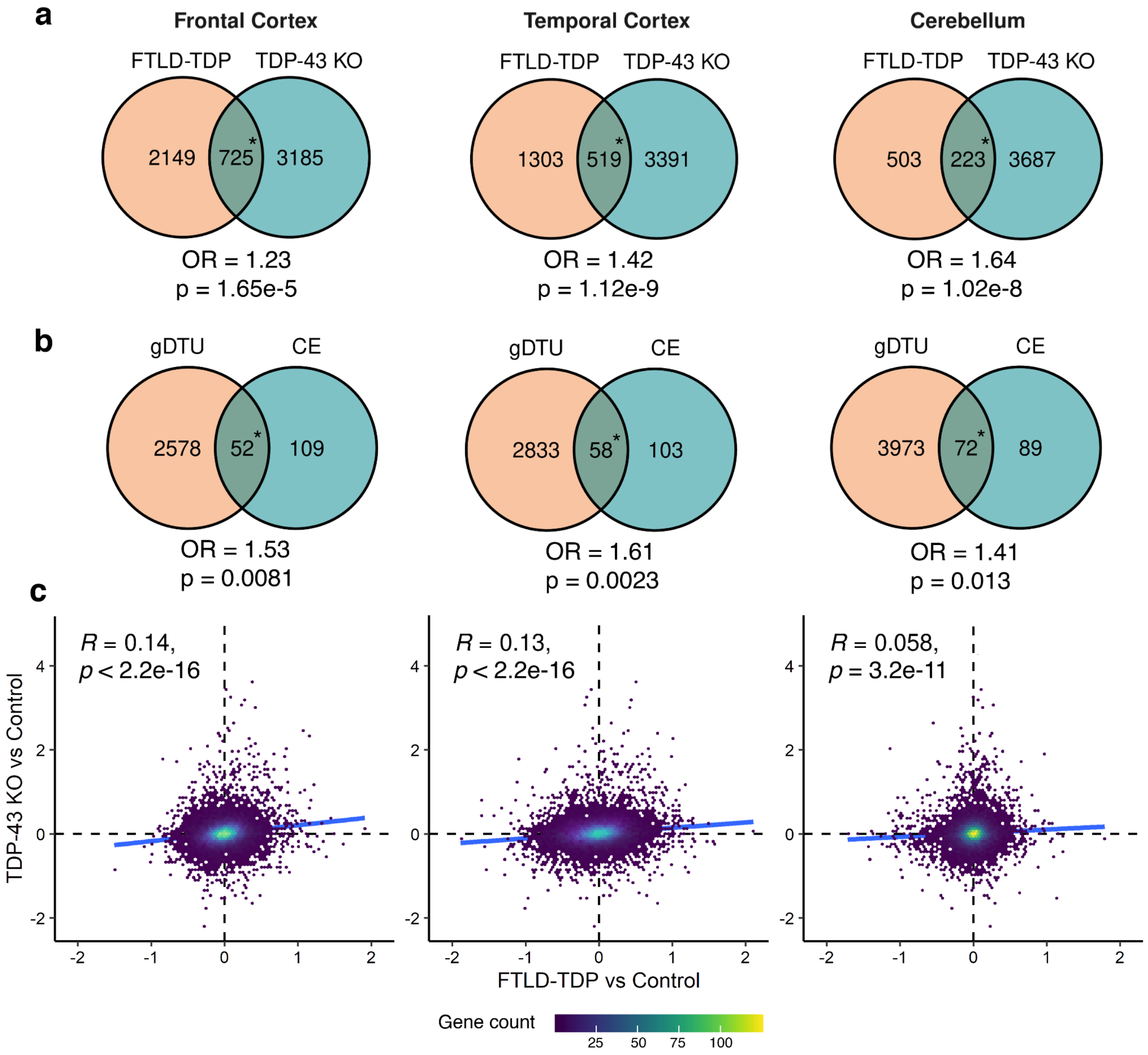


**Supplementary Figure 18: Comparisons with TDP-43 knockdown genes from Brown et al.**

(a) Comparing differentially expressed genes between FTLD-TDP vs Control and TDP-43 knockdown (Brown et al, 2020). (b) Comparing DTU genes (gDTUs) with genes with possible cryptic exon (CE) inclusions (Brown et al, 2020). Odds ratios (OR) and P-values for all overlaps were computed using a one-sided Fisher exact test. (c) Scatter plots show correlation between $\log_2$ fold changes per change between the FTLD-TDP samples and controls (x axis) and TDP-43 knockdown and control cell lines (y axis). Individual points are genes, color refers to density of overlapping points. R refers to the Spearman correlation coefficient.
